# MACE prediction using high-dimensional machine learning and mechanistic interpretation: A longitudinal cohort study in US veterans

**DOI:** 10.1101/2022.10.31.22281742

**Authors:** Sayera Dhaubhadel, Beauty Kolade, Ruy M. Ribeiro, Kumkum Ganguly, Nicolas W. Hengartner, Tanmoy Bhattacharya, Judith D. Cohn, Khushbu Agarwal, Kelly Cho, Lauren Costa, Yuk-Lam Ho, Allison E. Murata, Glen H. Murata, Jason L. Vassy, Daniel C. Posner, J. Michael Gaziano, Yan V. Sun, Peter W. Wilson, Ravi Madduri, Amy C. Justice, Phil Tsao, Christopher J. O’Donnell, Scott Damrauer, Benjamin H. McMahon

## Abstract

High dimensional predictive models of Major Adverse Cardiac Events (MACE), which includes heart attack (AMI), stroke, and death caused by cardiovascular disease (CVD), were built using four longitudinal cohorts of Veterans Administration (VA) patients created from VA medical records. We considered 247 variables / risk factors measured across 7.5 years for millions of patients in order to compare predictions for the first reported MACE event using six distinct modelling methodologies. The best-performing methodology varied across the four cohorts. Model coefficients related to disease pathophysiology and treatment were relatively constant across cohorts, while coefficients dependent upon the confounding variables of age and healthcare utilization varied considerably across cohorts. In particular, models trained on a retrospective case-control (Rcc) cohort (where controls are matched to cases by date of birth cohort and overall level of healthcare utilization) emphasize variables describing pathophysiology and treatment, while predictions based on the cohort of all active patients at the start of 2017 (C-17) rely much more on age and variables reflecting healthcare utilization. In consequence, directly using an Rcc-trained model to evaluate the C-17 cohort resulted in poor performance (C-statistic = 0.65). However, a simple reoptimization of model dependence on age, demographics, and five other variables improved the C-statistic to 0.74, nearly matching the 0.76 obtained on C-17 by a C-17-trained model. Dependence of MACE risk on biomarkers for hypertension, cholesterol, diabetes, body mass index, and renal function in our models was consistent with the literature. At the same time, including medications and procedures provided important indications of both disease severity and the level of treatment. More detailed study designs will be required to disentangle these effects.

## Introduction

Cardiovascular disease (CVD) is implicated in ∼ 25% of deaths, both in the US [2] and worldwide, where it was responsible for 18.6 million deaths in 2019 [65]. Epidemiological studies of CVD consistently find that risk is associated with non-modifiable risk factors such as age and sex, with biomarkers for low-density lipoprotein (LDL) cholesterol, hypertension, and diabetes, and modifiable risk factors such as obesity, smoking, and lack of physical activity [81, 16, 33, 26, 65]. However, there is substantial heterogeneity in how these risk factors impact patients’ morbidity and mortality. The co-occurrence and interdependence of many of these pathophysiological CVD risk factors is defined as a metabolic syndrome [22]. A better understanding of the impact of diverse risk factors in CVD will have important implications, given the large number of associated clinical issues and healthcare burden [41].

Advances in artificial intelligence (AI) have led to a variety of applications in analyses of electronic medical records (EMR) [69], with detailed neural network architectures proposed to capture the logic of EMR analysis [61]. In recent years, several studies have assessed the value of AI in predicting outcomes for chronic diseases. Singh *et al*.[70] predicted deterioration of renal function in a cohort of 6,435 patients with a variety of longitudinal AI models, concluding that longitudinal, multi-task predictors showed great promise, yet they emphasized that the details of problem formulation significantly impacted results. Zhao, *et al*. [82] compared multiple AI techniques applied to several data sets in analyses of 109,490 CVD patients, and concluded there was minimal accuracy improvement with the AI techniques, but some benefit to larger sets of predictor variables, compared to the American College of Cardiology (ACC) and the American Heart Association (AHA) guidelines. [33]. Steele, *et al*. [73] predicted coronary artery disease mortality in a cohort of 80,000 patients, concluding that AI models worked well without imputation, that random forest models did not outperform Cox models, and that variable selection techniques could identify important, novel, predictors in a data driven manner.

Many of the challenges in making practical use of AI for medical outcomes prediction could be anticipated from the general difficulties associated with deep learning methodologies, as described by Marcus [51]. In particular, these methods are data hungry, opaque to feature importance, difficult to integrate with prior knowledge, and work poorly with hierarchical structures. They also assume stable relationships among variables and thus do not support robust engineering. Sculley, *et al*. [68] argued that these aspects of deep learning models lead to substantial hidden costs in deploying deep learning solutions to practical problems, as they are brittle and often need to be re-trained with fairly innocuous changes in details of the input data stream.

The need for robustness in solving medical outcomes problems has led to a variety of detailed comparisons of logistic regression, Kaplan-Meier analysis, and Cox proportional hazard models [23, 16, 55]. Notably, the critique that retrospective studies are intrinsically less reliable than randomized, controlled trials may depend significantly on our ability to define predictor variables in a way that minimizes bias [10, 9]. Another difficulty with traditional regression models is that they often neglect to incorporate known interaction terms, creating misleading associations with an outcome [78]. Thus, it is important to assess not only the calibration and subgroup performance of predictive models, as described in Ref. [40], but also to construct models in a more generalized way to be applicable beyond the specific study design and population used to create the models [39]. Note that there are several classes of methods used to probe the determinants of classification in deep learning, as reviewed in Ref. [46]; we confine ourselves here to interpretation of logistic regression coefficients.

In this work, we predict MACE events in millions of veterans based on analyses of hundreds of longitudinal, hand-curated variables from the VA electronic medical records (EMRs) describing demographics, diagnoses, laboratory results, medications, and treatments, with 7.5 years of predictor variables and follow-up of up to 15 years. We begin by describing the selection, cleaning, and encoding of data in our chosen study designs, which include both retrospective case-control (Rcc) and prospective designs. This is followed by characterization of model accuracy for several prediction methodologies, including Cox models and interval prediction with logistic regression and LASSO model selection, as well as three machine learning (ML) approaches - a simple neural network (Nnet), a neural network with an attention scheme (Tabnet), and a random forest (RF). Included in our analyses is the transfer learning across study designs, based on a re-optimization of selected variables to account for specific differences among the study designs. For the case of logistic regression on Rcc, we examine in some detail predictor variables relating to disease pathophysiology. Finally, we compare model coefficients for Rcc and the cohort of all active patients at the start of 2017 (C-17), analyze associations of Rcc model variables with individual components of our combined outcome, and examine the dependence of selected variables on two key confounding variables, age and the level of healthcare utilization.

## Methods

### MACE outcomes and study designs

Our outcome (and its time) was defined as the first recorded instance (and that time) of any one of three types of MACE (Major Adverse Cardiac Events): acute myocardial infarction (AMI), ischemic stroke, and ASCVD death. For AMI and stroke, we used International Classification of Diseases (ICD-9 and ICD-10) codes from inpatient and outpatient records, and hospital service fees. For ASCVD death, we linked our patient data to the underlying cause of death in the National Death Index (NDI) database. For AMI, both 410.x (acute myocardial infarction) and 412.x (old myocardial infarction) were used for ICD-9 codes, while I21.x (acute myocardial infarction), and I22.x, and I25.2 (old myocardial infractions) were used for ICD-10. For stroke, 433.[012389]1 (occlusion and stenosis of cerebral arteries, with infarction, where square brackets indicate any of the enclosed digits can be matched) and 434.91 (occlusion of cerebral arteries with unspecified infarction) were used for ICD-9 codes, while I63.x (cerebral infarction) were used for ICD-10. In addition to the AMI and stroke conditions, the cause of death file was searched for any I25, (chronic ischemic heart disease), Z95.1 (presence of aortocoronary bypass graft), and Z98.61 (presence of coronary angioplasty implant and graft) codes.

We define ‘time of prediction’ as the date when prediction of a given outcome, which occurs later, is made. For all study designs (see Figure 1), any record of the above-defined outcomes prior to the time of prediction resulted in exclusion of the patient as either a case or a control. We analyzed four cohorts of patients each with its own additional criteria. For the Rcc cohort, the ‘time of prediction’ was required to be after July 1, 2007. For the LDL decision support (LDL-DS) cohort, at least one cholesterol lab reported in the 3 yrs prior to July 1, 2010 was required, and the time of prediction was one week after the first such result. Similarly, for the visit decision support (Visit-DS) cohort, at least one outpatient visit was required in the 3 yrs prior to July 1, 2010, and the time of prediction was one week after the first such visit. Finally, for the C-17 cohort, at least one inpatient or outpatient visit was required in the three months prior to the ‘time of prediction’, which was January 1, 2017 for all patients in this cohort.

**Figure 1:**
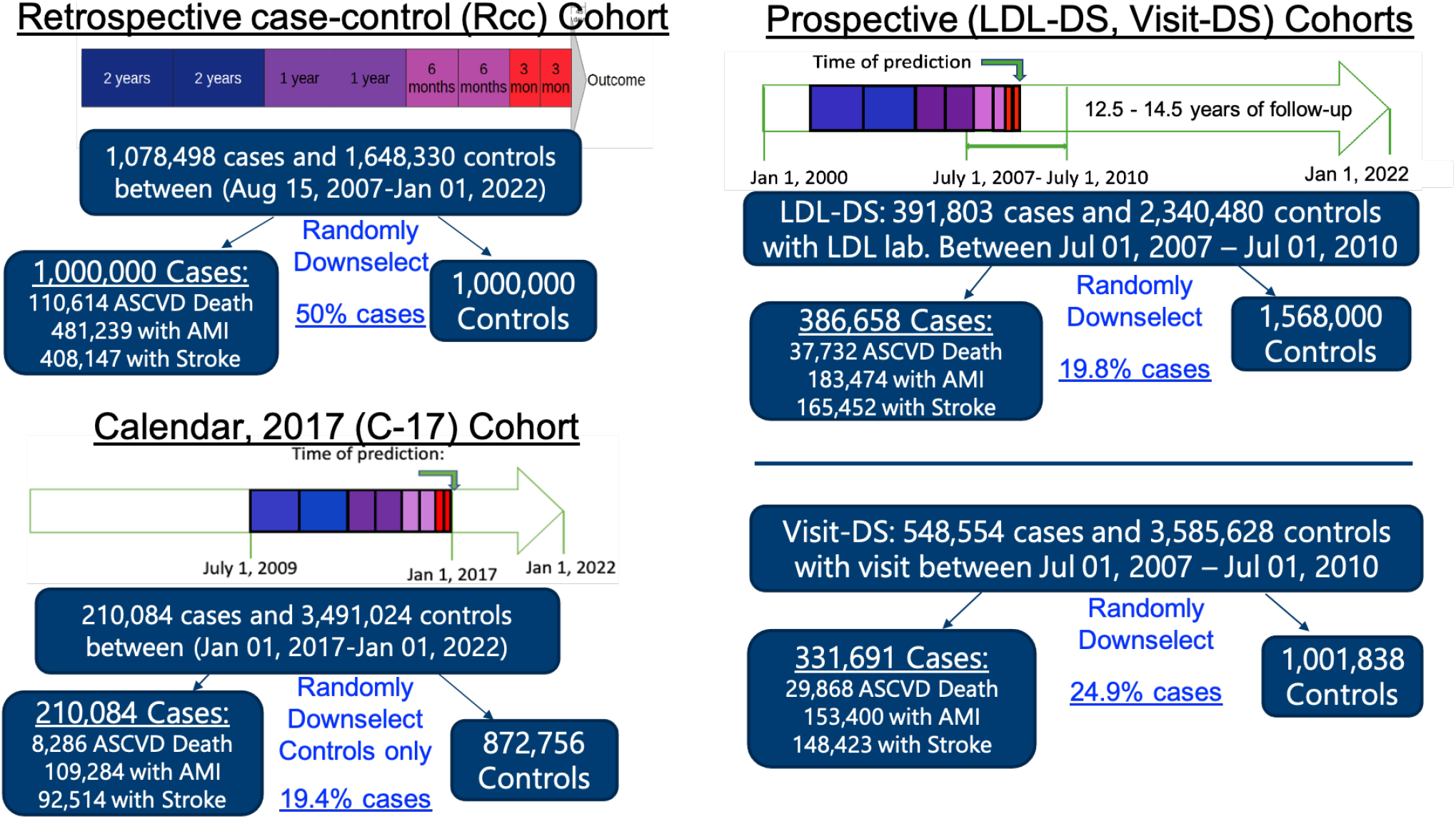
The four study designs used in this work. **(top left)** The retrospective case-control (Rcc) study aggregates predictor variables in eight time windows of data collection, ranging from 45 days to 7.5 years prior to a MACE outcome. The outcome is either a first recorded MACE event after August 15, 2007 and before January 1, 2022 (cases) or a visit chosen randomly from a list of visits during that time period from the same date-of-birth cohort (controls). Patients with a recorded MACE event prior to August 15, 2007 were excluded from being either cases or controls. **(right)** Two prospective cohorts were defined for time-to-event (Cox) models to mimic decision support conditions. For both, the time of prediction was defined to be one week after, respectively, the first return of a LDL cholesterol measurement (LDL-DS) or the first visit for any purpose (Visit-DS), occurring between July 1, 2007 and July 1, 2010. Follow up was until January 1, 2022. Predictor variables were aggregated into 8 time windows spanning 7.5 years before the time of prediction in a manner identical to that used for Rcc. **(bottom left)** Our C-17 cohort was defined with calendar dates, with all patients with a visit between October 1, 2016 and January 1, 2017, and no previous MACE event, included. The prediction was made on January 1, 2017, and predictor variables were aggregated into time windows as in the other cohorts. Details of the study designs are provided in the Methods section and attributes of the study populations are provided throughout the Results section.

Additionally for all designs: 1) we required a date of birth post-January 1, 1901 and a registration with the VA by Jan 1, 2022, when the data tables were created, and 2) we included a 4-6 week gap between the ‘time of prediction’ and the time of the first ‘allowed’ outcome, in order to minimize predictor variables from ‘leaking’ information from the very same visit defining the outcome. A very small number (dozens) of patients were excluded on the basis of failed self-consistency checks, such as recorded dates of death in the future, perhaps resulting from inconsistent information when patients registered at multiple VA Medical Centers.

Our Rcc design used as cases all first MACE events occurring between Aug 15th, 2007 and Jan 1st, 2022, and prediction was made with data spanning 7.5 years before the event. Controls were selected by randomly choosing visits of patients among non-cases during the same time period as the matching outcome event and from the same date of birth cohort. This study design allows the greatest amount of information (both chronic and acute) to be used, utilizes all of the available cases, removes age as a confounding variable, and balances overall healthcare utilization between cases and controls, potentially providing more insight into physiology and mechanism of disease. In this study design, we have 1 million cases and 1 million controls (Figure 1). For matching in the Rcc cohort, two criteria were used. First, controls were chosen from within one of 100 equally sized and time ordered date-of-birth cohorts of roughly 230,000 patients each. Second, in order to balance the average healthcare utilization of controls and cases, controls were chosen randomly among visits of patients who were not excluded or cases. This approach increases the likelihood of a patient being chosen as a control in proportion to the number of its visits. Our second study design, LDL-DS, was designed as a time to event model with an opportunity for at least 12.5 years of follow up, and the time of prediction set one week after LDL cholesterol labs were obtained. The LDL labs were required to be obtained during the three year period from July 1, 2007 to July 1, 2010. No matching of cases to controls occurred, so there were approximately 40% as many cases, but nearly twice the controls as in Rcc. Our third study design, Visit-DS was similar to LDL-DS, but required only a visit, rather than an LDL lab. Consequently, there were nearly twice as many of both cases and controls, compared to LDL-DS.

Finally, we created a calendar study design, C-17, that predicted MACE events on January 1, 2017 for the five year interval after February 8, 2017, for patients with a visit during the three month period before the time of prediction. This design has the lowest fraction of cases (5.7%) but reflects a recent sampling of patients utilizing the VHA system, and we analyzed it, together with Rcc, in most detail. To simplify the calculation and improve the machine learning model performance, we down-sampled the controls by a factor of four, computed randomly within each date-of-birth cohort. This kept the enrichment of cases in our actual predictive model calculations within a factor of two of each other (from 24% to 50%), which is helpful when comparing results between models.

Our predictor variables include patient age, gender, race, ethnicity, and 243 time-dependent, binary variables, which include vitals signs, lab results, diagnosis, procedure codes, and pharmacy fills data. For age, gender, race, and ethnicity, many patients were queried multiple times. In this case, the most frequent (mode) of the answers was retained. Gender was encoded as “male” (reference) and “not male”. Race was encoded as Black or African American or “not” (reference), and ethnicity was encoded as Hispanic (reference) or “not”.

Our time-dependent predictor variables came from a number of data domains, including demographics, ICD-9 and ICD-10 diagnosis codes, medication generic names, procedure codes (Current Procedural Terminology - CPT), vital signs, hospital resources utilized (stop codes) and laboratory results. The time dependence was organized backwards from the time of prediction in adjacent periods two each of 3 months, 6 months, 1 year, and 2 years, for a total of 7.5 years of data, as illustrated in Figure 1. This was in an effort to balance the need for parsimony against the desire for acute information while retaining 7.5 years of data. We refer to these periods as the time windows of data collection.

### Medications, procedures, and diagnoses

The drugs and drug categories for: antidepressants, stimulants, opioid-for-pain, estradiol, levothyroxine, ezetimibe, bile acid sequestrants (BAS), aspirin, warfarin, and metformin were defined by searching the “DrugNameWithoutDose” field for a list of words or word stems, and manually checking the output for relevance. The statins were mapped into low, medium, and high intensity based on the drug name and dosage following Grundy *et al*.[34]. Cardiovascular drugs and diabetes drugs were identified by using the “VADrugClassification” scheme, such as HS502 for oral hypoglycemic agents, HS501 for insulin, and most of the categories of CV drugs: CV050 for digitalis glycosides, CV150 for alpha blockers, CV200 for calcium channel blockers, etc. The VA drug classification scheme can be found online at, for example, [77]. We also created a category “Any Drug” simply matching any of the list of all products distributed by the pharmacy, including non-drug products such as lancets. A spreadsheet with the most common medications comprising each group is provided in the SI. Medications were recorded as present in each time window of data collection if one or more instance were recorded and absent if none of the medications included for each variable was present.

Procedure codes were selected from among the most prevalent inpatient and outpatient CPT codes identified as pertinent to CVD, grouped manually by inspection, and included EKG12 (9300, 9305, 93010, and 93225), rEKG (93040, 93041, and 93272), doppler (93307, 93320, and 93325), dialysis (90935, 90937, 90999, 90989, 36145, and 36489), stress test (9301[5678] and 93350), artery scan (93875, 93880, and 9392[2346]), veinous study (93965, and 9397[0610]), ESRD service (9092[15]), left heart catheter (93510), coronary angiography (93539, 9354[035], and 9355[56]), pacemaker (9373[123]), cardiac rehabilitation (9379[78]), lung capacity test (93720), blood pressure monitoring (93784 and 93790), stent (92980), and defibrilator (93741), where square brackets indicate any of the enclosed digits can be matched. Procedure codes were recorded as present in each time window of data collection if one or more instances were recorded and absent if none of the codes included for each variable was noted.

Variables defined from ICD-9 and ICD-10 codes were adapted from a list provided by the VA Office of Mental Health and Suicide Prevention, following from definitions used in Ref. [52]. Diagnosis codes were collected from inpatient, outpatient, and fee-for-service tables. ICD-9 and ICD-10 codes were lumped together, with letters for ICD-9 codes (used before October 1, 2015) changed to lower case and all search terms required to match from the first character in the field. For example, using this process, alcohol use disorder is characterized by the variable alcdx_poss and matches any of ICD-9 codes starting with 291, 303, and 305.0 and ICD-10 codes starting with F10. Variables are sometimes overlapping in the codes they use and were included with the idea that Lasso model selection would identify the best overall for the MACE prediction problem. An important aspect is that variable definitions should ideally be consistent over time, including the ICD-9 to ICD-10 changeover in 2015, and also through the yearly revisions of the coding manuals. To this end, we plotted histograms (see the SI) of Dx-based variables together with the number of occurrences of the most prevalent component variables for a 3% sampling of representative aged patients. This allowed verification that there was consistent reporting through time, and we attempted to harmonize the definitions as much as possible, but a certain degree of confounding is likely to occur. Diagnosis-based variables were recorded as present in each time window of data collection if one or more instances were recorded and absent if none of the codes included for each variable was noted.

### Vital signs and laboratory data

Vital signs were taken from both inpatient and outpatient visits, and included systolic and diastolic blood pressure, weight, pulse, respiration rate, oxygen saturation and self-reported pain (scale from 1 to 10). Laboratory results were collected from the “LabChemTestResultNumeric” field. For each of these variables, the median reported value was computed for each patient during each of the eight time windows of data collection. For all vital signs except pain, and all laboratory results, values were imputed for time windows without recorded values by first carrying values forward in time to replace NAs, then backwards in time to replace any remaining NAs. The reasoning is that recorded values are observations of a continuous process; as such the most recent observation should be used in the case of missing information. By contrast, reported pain, diagnoses, procedures, and medications are considered events, and the presumption is that when they are not noted it is because they did not occur. Weight was transformed into BMI using the median height for each patient, computed from all recorded measurements of height for each individual patient, and any values below 10 or above 100 were discarded.

Laboratory results and vital signs were binned according to standard clinical nomenclature and referenced to the ‘normal’ category. Note that lack of measurement is included as an ‘NA’ category, which is potentially informative. Specifically, BMI (*kg/m*^2^) was classified as (i) underweight if < 18.5, (ii) overweight if > 25 and < 30, and (iii) obese if > 30, with the reference state, ‘normal’, being between 18.5 and 25. Similarly, for systolic blood pressure (*mmHg*), (i) hypotension is < 90, (ii) elevated blood pressure is between 130 and 140, (iii) hypertension 1 is between 130 and 140, and (iv) hypertension 2 is greater than 140; with the reference state, ‘normal’, between 90 and 130. A similar process was used for the other vital signs. Pulse pressure was computed by subtracting reported median diastolic from reported median systolic, for a given time period. Because of the way results were carried forwards and backwards across time bins, patients in the NA category will have NA recorded for all eight time windows of data collection.

Laboratory results in the VA medical records are assigned an unique identifier (“LabChemTestSID”), along with a location (“sta3n”) and sample type (“topology”) codes. On the basis of these fields, we have established lists of identifiers associated with 100 of the most common laboratory result types, and we selected a subset of these labs for use as MACE predictor variables. Laboratory results were processed in a similar manner to vital signs, except that all labs had an NA category included, so a patient not identified as NA, high, or low is a patient whose median result for that time window is within the normal range for that lab result. Second, histograms of median results across patients were inspected for each laboratory result type, and cutoff values were identified as likely separating values using other than those expected; values outside the ‘reasonable’ range were assigned to the NA category, as with missing results. For the case of creatinine, values >10 were divided by 88.2, to convert from *µ*mol/L to g/dL before out of range values were identified, and the standard gender-dependent and race-dependent formula was used to convert to eGFR. If both eGFR and creatinine were reported in a given time interval, the eGFR value was used. For hemoglobin levels and red blood cell counts, distinct mappings between values and categories were used for men and women, with 11-12 g/dL assigned ‘low’ for men, and 10-11 g/dL assigned ‘low’ for women for hemoglobin level, and the ‘normal’ range for RBC was between 4.5 and 5.5 for men and 3.8 and 4.8 for women. For differential WBC, no attempt was made to process absolute measures of WBC categories; if values were not in the range expected for percentage, they were assigned to the NA category.

### Prediction methodologies

Six different modeling algorithms were used to predict the occurrence of a MACE event over the time window of prediction, given the 7.5 years of medical history of the patients before the time of prediction. The algorithms used are i) a AHA/PCE [33] like equation for baseline, ii) logistic regression (GLM) [13, 23] with LASSO [76] penalty for model selection [17], iii) random forests (RF) [5], iv) a fully connected (vanilla) neural network (NN) [20, 35, 53] with 3 hidden layers, v) a transformer based neural network (TabNet) [4], and vi) a Cox model [12].

We recreated the American Heart Association’s Pooled Cohort Equations [33] (AHA-PCE) prediction model by matching each of the predictor variables in that model, as best as possible, with the variables in our data set containing the 247 predictor variables. Demographics information were taken as is from our data. Information on the cholesterol level was extracted from the HDL cholesterol, LDL cholesterol, and total cholesterol variables and encoded as present if it was present in any of the eight time bins for the three variables. For smoking, the Nicdx variable which includes the diagnosis codes for nicotine use (ICD9 305.1, and ICD10 F12.2 and Z72.0) was used. Blood pressure information was extracted from the hypotension, elevated blood pressure, hypertension_1 and hypertension_2 variables. To encode whether or not a patient is on blood pressure medications, a combination of ace-inhibitors, angiotensin receptor blockers, anti-anginals, alpha blockers, beta blockers, calcium channel blockers, loop diuretics, thiazide diuretics, and potassium sparing diuretics variables was used and encoded as a yes if the patient was on any of these medications in any of the eight time bins. To encode whether a patient had diabetes, a combination of diagnosis of diabetes, labs that show elevated glucose, or hemoglobin A1C greater than 7%, and prescription of insulin or other diabetes drugs such as sensitizers, metformin, or thiazolidinediones was used.

For all methods except Cox, the problem was formulated as an interval prediction problem with a binary outcome. This design was applied to the Rcc and C-17 cohorts, and analyzed with AHA/PCE-like model and logistic regression, as well as machine learning algorithms - random forest, simple fully connected neural network, and TabNet. The logistic regression model was tuned with the Lasso penalty to enable model selection of the predictor variables. For random forest, we used 500 trees, the square root of the number of input features for the number of features to consider for the best split, and require a minimum of 100 samples at a leaf node. The fully connected neural networks had 3 hidden layers with 1024, 512, and 256 neurons respectively and ReLu non-linearity [27]. For Tabnet, the Pytorch implementation was used [19] with default settings for the hyperparameters, including 8 for the width of the decision prediction layer, 8 for the width of attention embedding, 3 for number of steps in the architecture, and 1.3 for gamma, which is the coefficient for feature reusage in the masks.

For Cox models, the study was formulated as a time-to-event problem. The LDL-DS and Visit-DS cohorts were analyzed with a Cox regression time-to-event approach. To facilitate comparison with the predictions of the other algorithms, after fitting the Cox models, they were evaluated as interval prediction at the indicated times-to-event. The LDL-DS and Visit-DS cohorts were also analyzed with AHA/PCE-like model by formulating them as an interval prediction problem. We calculated c-statistics (or area under the receiver operating characteristic - ROC - curve) to compare the different models and study designs. As part of this comparison, we evaluated the c-statistic both with the trained model used directly on each study design, and with a version of the trained model ‘re-optimized’ by computing a logistic regression (GLM) model on a subset of variables, including the model score described, together with Gender, Race, Ethnicity, Age, Age^2^, Pain > 0, AnyDrug, HbA1c > 9%, DEMENTIA, and EH HYPERTENSION.

For all study designs and methodologies, we use half of the cases and controls to train the model, and the other half to evaluate it.

## Results

Table 1 compares subject characteristics across the four cohorts of patients we created. Motivation and detailed descriptions for each are provided in the Methods section and Figure 1. We have between 210,084 and 1,078,498 cases across study designs (top row). Because statistical power was not an issue with our large numbers of cases and controls, and because several of the prediction methodologies perform better with balanced data sets, we down-sampled the cohorts for calculation, as indicated in the bottom row of Table 1. As is common in the VA Healthcare system, females, African-Americans and Hispanics have age distributions that differ from the overall average, which is dominated by white males. Details characterizing these distributions and the raw breakdown of cases across sub-outcomes (AMI, stroke, and ASCVD death) and demographics are provided in the table.

**Table 1:** Selected characteristics of each of the four study populations, broken down by cases and controls. The first row provides the total number of cases and controls in each study, together with the percentage of cases in each group, which drops from 39.6 % in the enriched Rcc study, to the 5.7% in C-17, which has only 5 years of followup. The next 3 rows provides the breakdown of the combined MACE outcome into AMI, stroke and ASCVD death. The breakdown of component outcomes is roughly 48:42:10 for all studies except C-17, which only has 4% ASCVD deaths, because our NDI data reporting ends after only 2 of the 5 years of followup, at the end of 2018. Below this are case and control breakdowns for three important and overlapping subgroups, females, Blacks, and Hispanics. From the reported percentage with MACE events for each group, we see that women have a much lower rate of events than overall and than each of the other subgroups. Some of this difference is determined by the age distribution, which are characterized in the bottom portion of the table.

|  | Rcc |  | LDL-DS |  | Visit-DS |  | C-17 |  |
| --- | --- | --- | --- | --- | --- | --- | --- | --- |
|  | Control | Cases | Control | Cases | Control | Cases | Control | Cases |
| **Total** | 1,648,330 | 1,078,498 (39.6%) | 2,340,480 | 391,803 (14.3%) | 3,585,628 | 548,554 (13.2%) | 3,491,024 | 210,084 (5.7%) |
| AMI |  | 519,225 |  | 186,274 |  | 255,752 |  | 109,284 |
| Stroke |  | 440,035 |  | 166,278 |  | 241,672 |  | 92,514 |
| ASCVD death |  | 119,238 |  | 38,058 |  | 51,120 |  | 8,286 |
| **Female** | 98,263 | 35,713 (26.7%) | 183,353 | 12,900 (6.6%) | 222,540 | 15,817 (6.6%) | 384,516 | 9,045 (2.1%) |
| **Black** | 264,330 | 163,829 (38.3%) | 368,813 | 66,450 (15.3%) | 511,674 | 87,694 (14.6%) | 629,516 | 40,916 (6.1%) |
| **Hispanic** | 133,052 | 48,670 (36.2%) | 133,052 | 22,673 (14.6%) | 175,052 | 25,684 (12.8%) | 56,892 | 11,818 (4.9%) |
| **Age (median)** | 69.3 | 69.8 | 60.1 | 63.1 | 62.7 | 64.6 | 64.2 | 69.2 |
| Female | 58.9 | 61.2 | 47.8 | 55.3 | 49.4 | 56.8 | 49.4 | 60.1 |
| Black | 63.7 | 64.7 | 53.9 | 59.0 | 55.5 | 60.5 | 58.1 | 65.2 |
| Hispanic | 66.8 | 69.3 | 57.6 | 64.0 | 58.5 | 64.3 | 56.7 | 69.2 |
| Youngest quartile | 56.1 | 56.9 | 42.2 | 47.7 | 44.8 | 50.4 | 40.2 | 47.9 |
| Oldest quartile | 84.9 | 85.2 | 79.6 | 79.0 | 81.0 | 80.5 | 79.9 | 80.1 |
| Computed dataset | 1,000,000 | 1,000,000 (50%) | 1,568,000 | 386,658 (19.8%) | 1,001,838 | 331,691 (24.9%) | 872,756 | 210,084 (19.4%) |

### Model Scores

Figure 2 and Table S1 present c-statistics for models trained with eleven combinations of prediction methodology and study design, and evaluated on the four study designs: Rcc, C-17, LDL-DS, and Visit-DS, as described in Table 1 and Methods. Figure 2 characterizes the discrimination power (c-statistic) of our predictive models. Each of the four panels shows scores for the evaluation test cohort indicated below the panel, but with models trained with eleven different combinations of study design and prediction methodology, as represented (and indicated) by each bar. The left most bar (green) has the lowest score in each panel, corresponding to our baseline AHA-PCE-like model trained on the panel’s test cohort. Next we present four different prediction methods - GLM, NN, RF, and TabNet - trained on Rcc; the Cox model trained on the LDL-DS and Visit-DS cohorts; and the same four prediction methods trained on the C-17 study design. Additionally, each model, with the exception of the AHA-PCE-like, was also ‘re-optimized’ on the study cohort being evaluated in each panel, with the improvement in score shown in red. The ‘re-optimization’ algorithm is based on a logistic regression with a subset of variables and the original model scores (see Methods).

**Figure 2:**
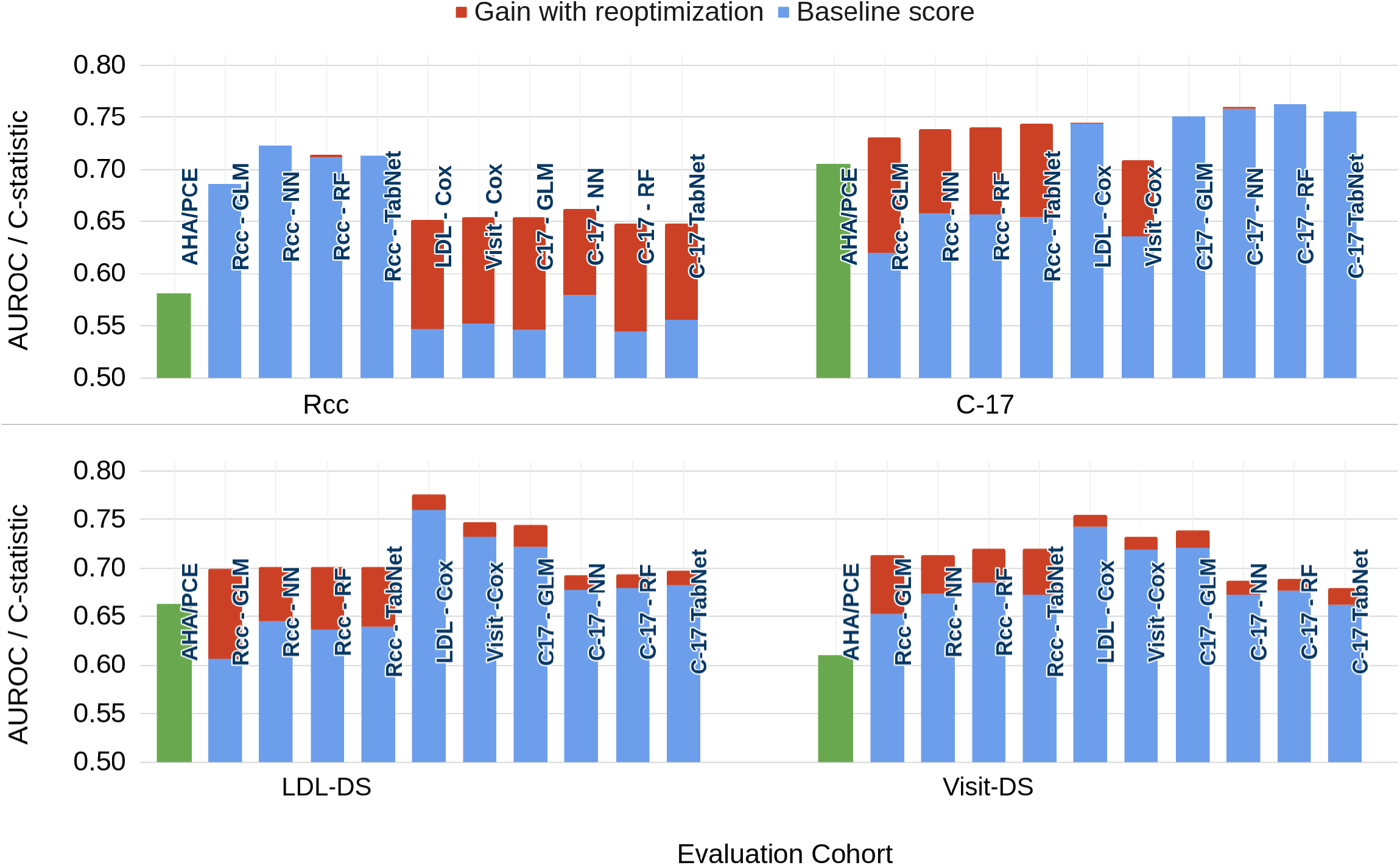
C-statistics for MACE prediction evaluated on each of the four study cohorts from Table 1, as indicated below each of the four panels. Models were trained on eleven prediction methodologies-study cohort combinations, as indicated with labels on each bar. The leftmost bar in each of the four graphs (green) indicates the c-statistic for our baseline model, the AHA-PCE-like model trained and evaluated on the indicated evaluation cohort. For the other ten bars in each group, the blue color represents the initial score of model with the indicated training and evaluation cohorts, while the red color represents the improved c-statistic when the model is re-optimized for the evaluation study cohort, as described in detail in the Methods section.

The scores for the pooled cohort equation (AHA-PCE-like) models vary considerably across the four study cohorts with the lowest score for Rcc cohort (∼ 0.58) and highest score (∼ 0.71) for the C-17 cohort. The low score for the Rcc cohort, which is loosely age-matched, may indicate the important role of age in the AHA-PCE-like equations. At the other extreme, the AHA-PCE-like equations, with their eight variables, performed nearly as well in C-17 as the re-optimized Visit-DS Cox model, with its 243 longitudinal variables.

Also notable is that the models trained on the three unmatched, prospective, cohorts performed equally poorly (∼ 0.55) when evaluated against the Rcc cohort, and substantially worse than the Rcc-trained models (all *>* 0.68). Even after re-optimization, their c-statistic was only 0.65. In contrast, models trained on the Rcc design are relatively good at predicting the C-17 cohort, with losses of only ∼ 2%, after re-optimization (red bars on top right panel of Figure 2), compared to C-17-trained models.

The two lower panels show model prediction scores re-optimized to 10-year interval predictions, in LDL-DS and Visit-DS cohorts. The LDL-DS cohort had a requirement of LDL lab data, rather than simply a visit, as is the case of Visit-DS. Eliminating the patients without the laboratory results produced better-performing models not only for evaluation on LDL-DS, but also Visit-DS (compare LDL-Cox to Visit-Cox bars in the lower right panel). Also striking is how much better the Cox models performed on LDL-DS than the interval prediction models.

### Model calibration in selected subgroups of patients

While c-statistics are commonly used to evaluate predictive models in medical outcomes analysis, they incompletely characterize model performance; calibration curves can provide additional insights [11]. Figure 3 shows calibration curves for four of our model calculations. In the calibration curves, the x-axis shows the model score and the y-axis is the observed probability of a MACE event, evaluated over different subsets of patients. In a well-calibrated model, all of the curves are given by the function shown as a solid black line in the figure (top row). In each of the four top panels, the black dots show the calibration curve for all of the patients in our test set (1M for Rcc, 0.5M for C-17), with symbol size proportional to the number of patients at each score. In the lower panels, we provide normalized histograms of the distribution across model scores, centered around a score of zero, which corresponds to a 50% probability of a MACE event. For all panels, colored dots represent the calibration curve and score histograms for distinct subgroups of patients, as defined in the figure legend and caption.

**Figure 3:**
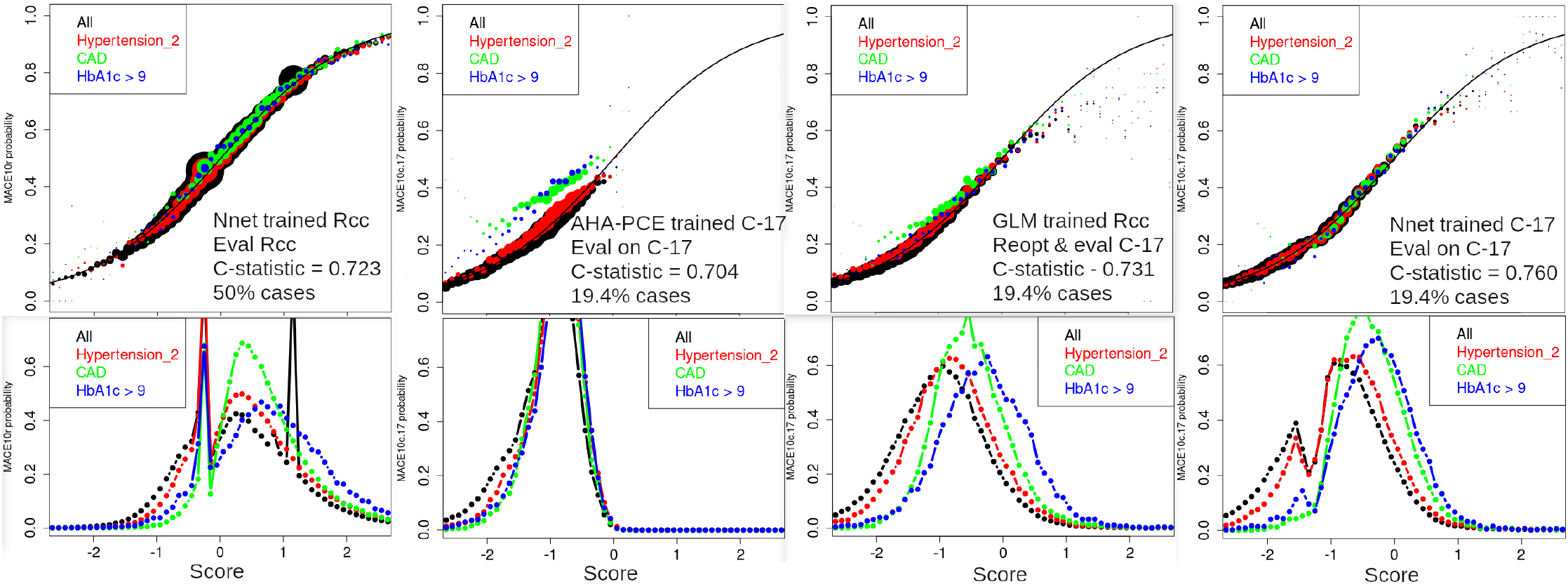
Calibration curves (top) and normalized histograms of scores (bottom) for all patients (black) and three subgroups of patients, defined by the presence of stage 2 hypertension (red), a diagnosis of coronary artery disease (green), and at least one time window with a median hemoglobin A1c ≥ 9% (blue). Symbol areas (top panels) are scaled to be proportional to the fraction of patients in each histogram with the indicated score. At the left are results from Rcc using the simple neural network, Second from the left to far right are results evaluated on the C-17 cohort, but trained in three different ways. First is our AHA-PCE-like model, next are results from our GLM re-optimized model, and at the far right are results from a neural network trained and evaluated on the C-17 data set. Neural network output probabilities, *Pr*, were transformed with the logit functionn, log(*pr/*(1 − *pr*)), before histograms were made, in order to facilitate comparison with logistic regression outputs.

The calibration curves in Figure 3 show results from four model calculations drawn from two cohorts of patients. In the leftmost panel of the figure, we present the simple neural network model (NN) trained and evaluated on the Rcc cohort of patients. Although most patients have predicted risks near 50%, as expected for a balanced data set, subgroups of patients are accurately placed at probabilities ranging from 5% to 95% probability of having a MACE event, and the curves for stage 2 hypertension, CAD, and HbA1c ≥ 9 also show excellent calibration (i.e., they follow the black solid line). Histograms of risk in the lower left panel show the model is able to identify high risk patients better than low risk. The sharp spikes in the histogram correspond to the risk scores for patients with only demographic information recorded; since Rcc is date-of-birth matched, the age-dependent risk does not spread these patients into distinct risk groups. The rightward displacement of the colored curves relative to black (for all patients) reflects the increased MACE probability associated with each of the defined subgroups.

For the other three panels, we compare the calibration of predictions from three different methodologies on the C-17 cohort, which includes 19.4% cases. As expected, the median probability of MACE event is now 19.4%, with the histograms of scores shifted to the left. Our baseline, AHA-PCE-like model (second column from the left in the figure), is unable to identify patients with MACE risk above 50%, and systematically under-estimates risk for patients with CAD and HbA1c ≥ 9, as expected for a model trained only on a few basic biomarkers. The third column shows our GLM trained on the Rcc cohort, then re-optimized for and evaluated in the C-17 cohort. The c-statistic improved by 3 percentage points, patients with MACE probabilities are accurately identified up to 60%, and the subgroups are much better-calibrated, although the risk for the low end risk of the CAD subgroup has their risk underestimated. As with the AHA-PCE model (second column), the risk histograms are all smooth, but in GLM model (third column), the increased risk associated with a CAD diagnosis and high HbA1c levels is clearly visible. The final model shown, a neural network model trained and evaluated on C-17 cohort, has the highest c-statistic (0.76), and it is also better calibrated across all the subgroups we analysed.

It is important to characterize model calibration for two reasons. First to verify that the model is accurately evaluating risk for particular variables, such as the risk factors shown in Figure 3. Second, to see that risk for important subgroups of patients is not systematically biased, which we address in supplementary information for gender, race, and ethnicity in Figure S3, age in Figure S4, and several subgroups which may impact healthcare utilization patterns in Figure S5. The most striking feature of Figure S3 is that all of the models are well calibrated across all four subgroups, and that females have significantly lower risk, even in the date-of-birth matched Rcc study. The pattern of spikes in the histograms of the Rcc neural net calculation suggest an interaction term between sex and healthcare utilization, but we do not attempt to disentangle this further in the present work. Careful examination of the curves show a small group (∼ 5%) of Blacks and Hispanics moved from medium (−0.8) to high (+0.8) risk, with a much smaller fraction in the age- and healthcare utilization-matched Rcc study. While the coefficient for Blacks in C-17 indicates a 14% higher MACE risk, that for Hispanics did not emerge through model selection.

Figure S4 breaks down the risk distribution by age for the four models. The three panels for the C-17 cohort all show relatively small differences for patients above age 60, and a one to two unit reduction in MACE score for the younger age groups. Reassuringly, the Rcc study, matched on date of birth, is able to accurately reproduce the lack of age dependence (see the histogram in the first column of Fig. S4), with good calibration throughout the risk curve. A systematic broadening of the width of the risk distributions for the younger age groups is evident, and the increased amplitude at low risk for the <50 group may be explained by the increased representation of females.

Figure S5 shows the risk distribution for four subgroups with distinct healthcare utilization patterns: those with diagnoses of homelessness, a mental health diagnosis, dementia, and a diagnosis of alcohol use disorder. The two regression models (second and third columns in the figure) is noticeably less well calibrated for every subgroup, with the largest difference for homelessness. The similarity of the score histograms in these subgroups to those for age, in Figure S4, is evident with the expected correlation of dementia in the > 80 and homelessness in the < 50 categories. Inspection of Table S2 also shows an association of homelessness with treatment: in the Rcc cohort, the 6.6% of patients with a homelessness diagnosis code reported have a 30% greater chance (than overall) of HbA1c > 9.0%, but a 10% lower chance of being prescribed insulin sensitizers. Whatever the origin of the relationship, it is evident that the NNet models are much better able to capture the relationship.

### Model Coefficients

Predictive model coefficients can provide insights into the correlates of MACE events, but we are faced with the question: which of the 84 calculations explored in Figure 2, or the four calculations described in detail in Figure 3 will be most informative? Model discrimination, presented in Table S1 and calibration curves and subgroup analyses presented in Figures 3 and S3 - S5 provide some guidance. We choose to focus on the GLM predictions using the Rcc study design because it maximizes the number of cases, averages over the past fifteen years, utilizes acute information for every case, and uses matching to minimize the impact of the confounding variables of age and level of healthcare utilization. We choose to examine the logistic regression calculation despite the substantial improvement in accuracy attainable with the other ML methods because the model assumptions of logistic regression are broadly understood.

We divide the predictor variables into two Figures. Figure 4 shows the coefficients for variables pertaining to metabolic syndrome, with laboratory results, diagnoses, and medication variables grouped together for each condition. In these stacked bar charts, the red colors at the base of each bar indicate the contribution to the coefficient of that variable for a time closer to the MACE event or control visit, over the 7.5 years of time of data collection corresponding to the redder shades in the arrow of Figure 1. In the bottom panel of Figure 4, the bars have a width proportional to the number of MACE cases with the given variable. Together with the detailed information provided in Table S2, and Figure S2, we can begin to understand much more about each predictor’s influence on MACE outcomes. We divide the variables pertaining to metabolic syndrome into four physiologically-related groups: BMI, hypertension and pulse rate, electrolytes and renal function, and lipids and diabetes.

**Figure 4:**
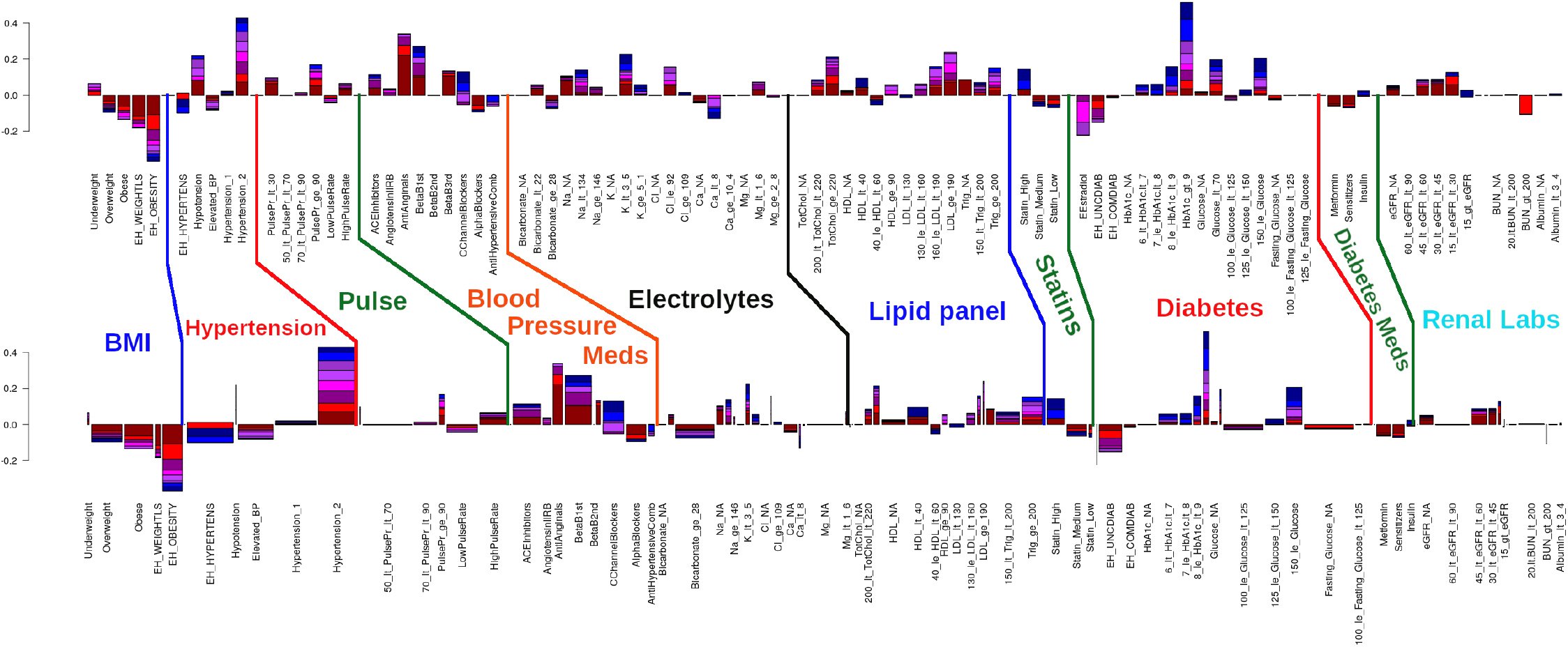
Stacked bar plots of our GLM model coefficients related to metabolic syndrome, from the Rcc cohort. (Top) Model coefficients, with the eight time windows of data collection stacked, with those proximal to the event at the base and colored red, while distal predictors (out to 7.5 years) are in bluer colors, corresponding to those shown in the arrow of Figure 1. Upward bars are predictive of a MACE event while downward bars indicate protection. Not shown are the reference categories for each lab and vital sign, which are taken to be the normal ranges. For most labs, lab_*N A* is included as a category indicating the patient did not have available data. (Bottom) The same data as at the top, but with the width of each bar proportional to the fraction of MACE cases in each category. The y-axis at left indicates the score resulting from the GLM model, and is in the range +/-0.4 for most of the variables shown. Analogous plots for C-17, LDL-DS, and Visit-DS, can be found in Supplemental Figures S6, S7 and S8

### BMI

Although BMI is a well-established risk factor for CVD [81, 16, 33, 26, 65], obesity is protective as a model coefficient (bars pointing down), whether characterized as a diagnosis code (EH_OBESITY; 278.0 or E66.x) or a vital sign, where overweight is defined as a median BMI between 25 and 30 kg/m^2^, while for obese, the median BMI is > 30.0 kg/m^2^. Although the coefficient for the Dx-code based variable is more than double that from the vital signs, it is important to recall from the Methods section that vital signs were imputed across time bins while the Dx-code based variables were not. This has the effect of increasing the impact of vital-sign-based coefficients in comparison to Dx- or medication-based. The widths in the bottom panel of Figure 4 are proportional to the number of patients with the code appearing at least once, and each time bin contributes separately.

Examination of the first few rows of Table S2 shows that BMI-related coefficients are much smaller (−0.08) for the C-17 study design, but also that the protective effect is not simply due to correlation with other risk factors, as the simple ratio of %cases/%controls also shows a protective effect. The dependence of raw percentage in Rcc and C-17 is evident in Figure S2 to be quite similar, at 32% and 41%, respectively. Also for both study designs, 85% of the patients with a Dx code for obesity have at least one record of BMI > 30, while 60% of those with a BMI > 30 have a diagnosis code of obesity.

### Hypertension and pulse rate

Stage 2 hypertension, defined as having a median systolic blood pressure measurement during a given time window greater than 140 mmHg, is one of the strongest MACE risk factors (Fig. 4), with a coefficient of 0.43 in Rcc and 0.53 in C-17 (Table S2), and a prevalence of 59% (Figure S2). Although the diagnostic code for stage 1 hypertension affects nearly as many people, its coefficient is neutral in Rcc and much smaller in C-19, at 0.15.

Each of the major classes of blood pressure medications are themselves strong and independent predictors of MACE events. Examination of the joint probability distributions in Figure S2 shows, however, that the likelihood of having a recorded pharmacy fill for blood pressure medications is only 30% elevated in subjects with stage 2 hypertension, perhaps indicating that the medications are effective in reducing blood pressure below 140 mmHg. While less prevalent than beta blockers, antianginal medications are both stronger MACE predictors and more acute (higher contribution of the dark red shaded area at the bottom of the bar in Fig. 4).

High pulse rate and pulse pressure are both predictive of MACE events, although high pulse pressure is only a strong predictor at higher levels (≥ 90 mmHg), where it is present in only 6.5% of patients.

### Electrolytes and renal function

Electrolyte imbalances, especially low sodium, potassium, and chloride are present in only a few percent of patients, but have coefficients in the 0.14-0.23 range in the Rcc study. High bicarbonate levels, which are associated with acid-base imbalance, are much more prevalent than the other imbalances and slightly protective.

Declining renal function, in each of three categories of eGFR between 60 and 15 ml/min/1.73m^2^ of body surface area, is acutely predictive of MACE events. The 30% of patients with eGFR < 60 ml/min/1.73m^2^ are 15% more likely to have stage 2 hypertension. From Figure S2, we see that they are also 50% more likely than the average patient to be taking first generation beta blockers.

### Lipid metabolism and diabetes

Low values of HDL (<40 mg/dL), high values of LDL (>130 mg/dL), high values of triglycerides (>200 mg/dL), and taking high intensity statins are all predictive of MACE events, with model coefficients of 0.1-0.2 and 5% to 25% of patients in each group. High intensity statin usage is 30% to 50% higher in patients with high LDL and triglycerides, compared to average, similar to the level of enrichment to that of a CAD diagnosis. The predictive value of the cholesterol labs is quite similar across study designs.

The contribution of diabetes risk to MACE events is multi-factorial and indicated by multiple diagnostic codes, laboratory results, and medications. Similar to the other categories of MACE risk factors, recently diagnosed diabetes and recently started diabetes treatments are protective, while abnormal laboratory results are predictive of poor outcome, with the highest levels (e.g., HbA1c) showing the strongest predictive coefficient.

As with the discussion of BMI, it is important to realize that laboratory results are imputed across all eight time bins, in contrast to both medications and diagnosis codes, which are only ‘present’ in time bins where they are reported. For diabetes, medication usage is acutely protective, while laboratory results are distal predictors of MACE events, with Dx codes in between. The protective nature of diagnoses and medications presumably indicates that attention is being paid to the condition, while the predictive nature of the laboratory results is because of their ability to more accurately indicate the severity of disease. For example, both Rcc and C-17 cohorts exhibit a much stronger risk for MACE when HbA1c ≥ 9% compared to lower values, similar to the highest category of glucose (>150 mg/dL) versus (125-150 mg/dL). When aggregated to indicate the top 16% of patients in terms of diabetes severity, HbA1c is about 20% more potent of a risk factor than glucose.

### Other predictors

Figure 5 is constructed in a manner similar to Figure 4, but characterizes predictor variables not directly related to the pathophysiology of metabolic syndrome, including specific CVD conditions and procedures, medications, mental health conditions, injuries and other laboratory assays. The strongest predictors in this figure are diagnoses of CAD or chest pains, with coefficients of 1.03 and 0.59, respectively. The most protective variable is a measure of healthcare utilization patterns: whether the patient had taken any drug during the 7.5 years prior to the time of prediction.

**Figure 5:**
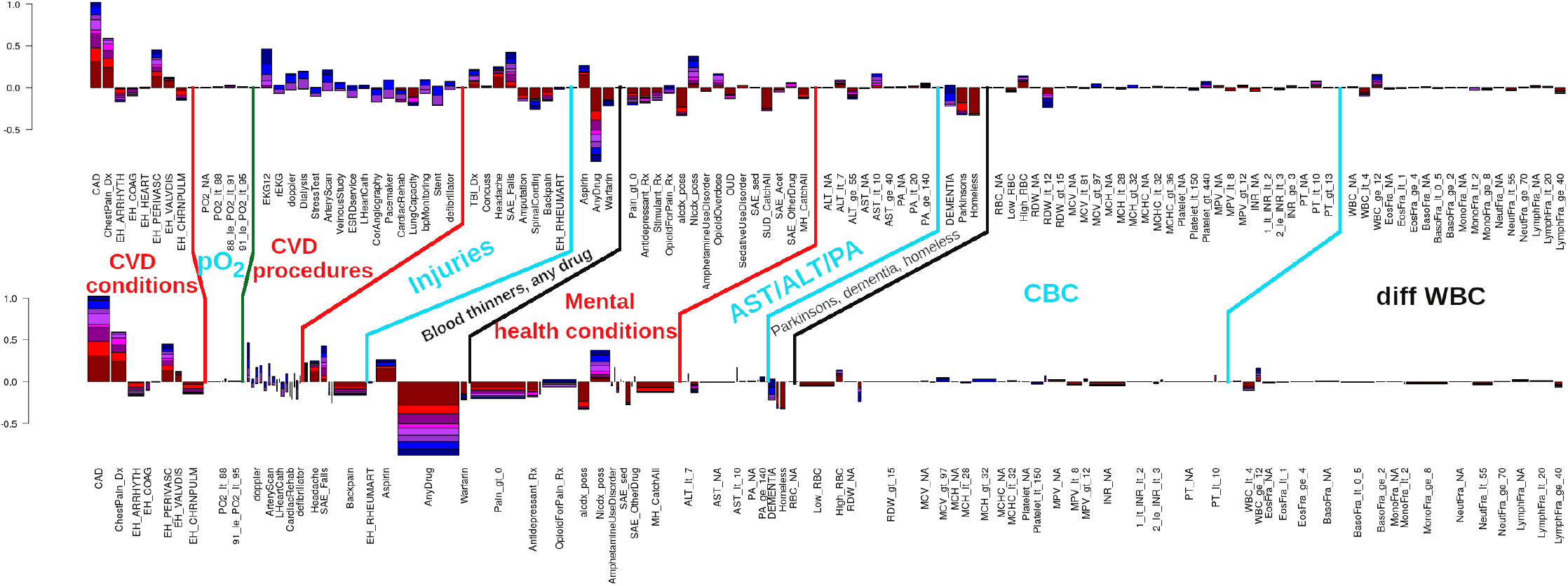
Stacked bar plots of GLM model coefficients related to CVD disease progression, comorbidities, mental health, complete blood count, and selected items from differential white blood counts, from the Rcc cohort. Panels and color-scheme for bars are as defined in Figure 4, except that the y-axis scale spans a broader range, to accommodate CAD and patients who have taken any drug. Analogous plots for LDL-DS, Visit-DS, and C-17 can be found in Supplemental Figures S6, S7 and S8

Analysis of complete blood counts (CBC) identified a number of statistically associations with MACE outcome, including liver enzymes, platelets, elevated white blood cell (WBC) counts, reb blood cell (RBC) counts, and red cell distribution width (RDW). These conditions are statistically significant, but comprise a much smaller population-attributable risk than the other variables discussed, and we consider them no further in this work.

Several mental health conditions correlate with MACE outcomes, and are generally protective. This may be because CVD and mental health care are two of the largest categories of VA care, and since we are not comparing to healthy controls, mental health is protective against MACE events. A notable contrast is nicotine dependence, which is strongly predictive of MACE events with a coefficient of ∼ 0.4 in all study designs, and with a prevalence > 20% in Rcc (see Table S2). A diagnosis of alcohol use disorder is protective (−0.33 in Rcc and neutral in C-17). While nicotine addiction is a well-known risk factor for MACE events, further investigation is required to understand whether the behavioral health codes are indicative of pathophysiology or healthcare utilization patterns. Aspirin is strongly predictive, presumably because it is prescribed to at-risk patients.

### Comparison across study designs, sub-outcomes, and confounders

While Figure 4 reproduces a variety of known risk factors with consistency across study designs, Figure 5 highlights the importance of predictors that are indicative of other effects, such as healthcare utilization patterns, age, or missingness of information. In order to better-understand these effects, we characterize how the results of models with our 247 variables depends on study design, components of our combined MACE outcome, and two confounding variables, age and healthcare utilization.

### Dependence on study design

The Rcc and C-17 study designs differ in many important aspects, with the most notable being the year-of-birth matching and enrichment of controls with healthcare utilization performed in the Rcc study. But they also differ in a variety of subgroup compositions, the time-period over which predictor variables are collected (i.e., always 7.5 years, but at different times prior to event in the two studies), the level of missingness of certain data types, and the relative importance of CVD onset vs. progression.

Figure 6 compares coefficients from the Rcc and C-17 study designs, with vital signs and laboratory results highlighted in the left panel and demographics, diagnoses, medications, and treatments highlighted in the right panel. In general, the vital sign and laboratory result predictors related to the pathophysiology as discussed with Figure 4 are highly correlated between the two study designs. This pattern includes important predictors such as stage two hypertension, HbA1c > 9%, and glucose > 150 mg/dl, as well as protective lab values for red cell distribution width (RDW), the liver enzyme ALT, and bicarbonate, and not having had a troponin test performed. Also evident in Figure 6 are a handful of variables that behave quite differently in the two study designs, such as reporting a non-zero level of pain (Pain gt 0), or not having had a reported respiration rate (RR_NA). We also observe a predominantly protective behavior from not having had laboratory results or vital signs measured at all during the 7.5 year window of prediction variables (green symbols). This may be due to the confounding effect that patients who utilize VA healthcare to a greater effect are more likely to have their AMI or stroke recorded, although a correlation caused by healthier patients not having heart attacks or CVD-related labs measured would produce a similar effect.

**Figure 6:**
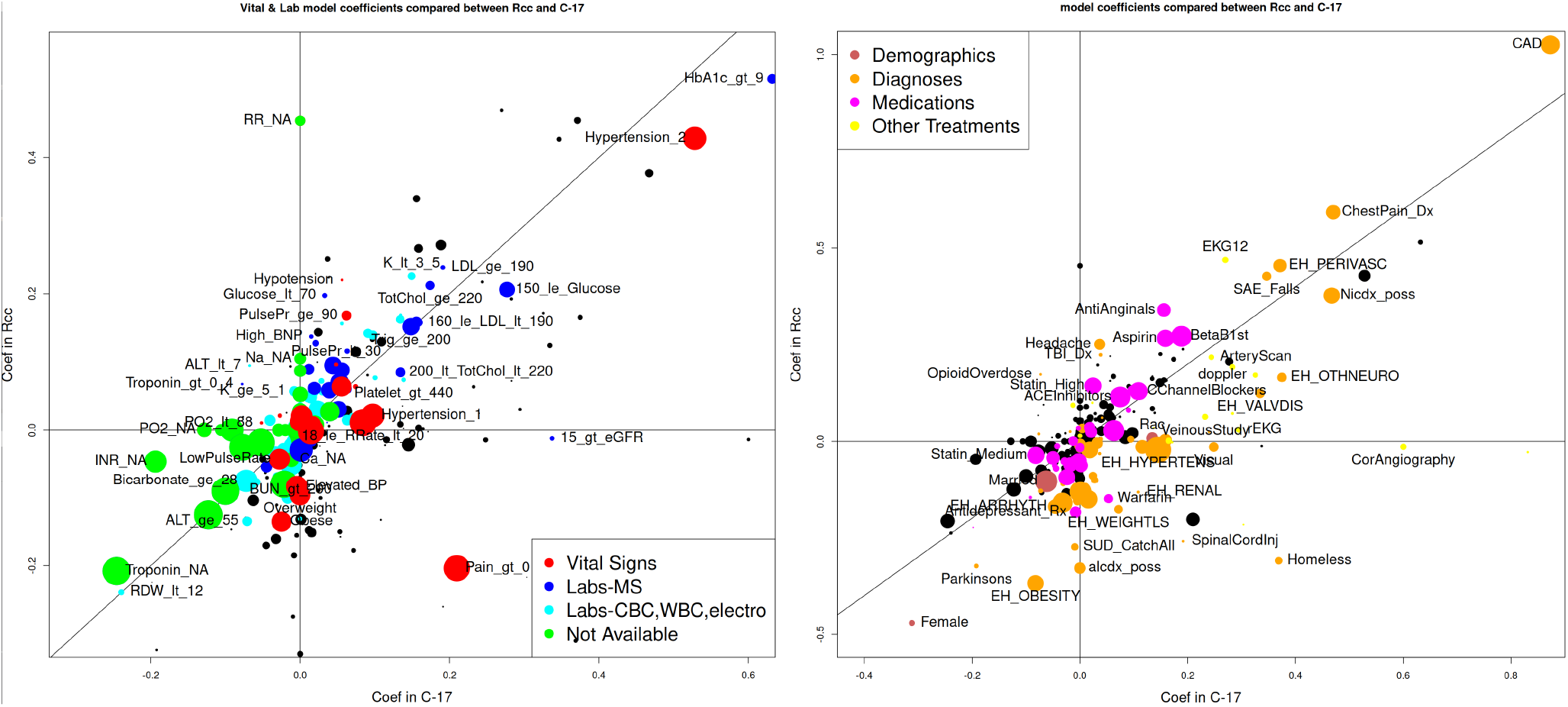
Comparison of GLM model coefficients between Rcc and C-17 study designs. The x-axis indicates the coefficient in the C-17 model, the y-axis indicates the coefficient in the Rcc study design, the symbol area is proportional to the number of cases with each attribute, and the color code as indicated in the legend. The left panel highlights variables associated with vital signs and laboratory results, while the right panel highlights demographics, diagnoses, medications, and other treatments. Variables of non-highlighted categories are provide in black, with reduced size, and selected variables are labeled. GLM coefficients and other information for all four study designs are provided in the SI. Horizontal and vertical axes and a line of slope one through the origin are provided for reference.

The right panel shows the same plot type, but for demographics, diagnoses, medications, and other treatments. It has an expanded scale, compared to the laboratory results, with generally good correlation between study designs, especially for the strong diagnosis-based (orange symbols) predictors of coronary artery disease (CAD), chest pains, nicotine dependency, peripheral vascular disease, and somewhat oddly, severe adverse effects from falls (SAE Falls). Notable exceptions to this trend include a diagnosis code for homelessness, alcohol and substance use disorders, and obesity. Women are protected from MACE events in both study designs, although more so in the Rcc than C-17. Medications are also correlated, but with much less dispersion than Dx codes.

Another clear effect that is visible in both panels of Figure 6 are groups of related codes with the same sign, but differing values of coefficients. Apparently, the models are grouping sets of partially overlapping parallel predictors in order to optimize overall scale. A variety of effects will contribute to this, such as differing statistical power of the study designs due to the enrichment of cases in Rcc (50%) compared to C-17 (19.4%), differing levels of disease severity, treatment patterns, and missingness between the two study designs. One large difference not visible in Figure 6 is the importance of age as a covariate in C-17, while it was matched for Rcc.

### Dependence on components of MACE outcome

Another important comparative metric to help understand the predictor variables is their level of enrichment in each component of our combined outcome. This is shown in Figure 7, using the Rcc study design and the same color scheme as in Figure 6. In this figure, we have plotted the level of enrichment of every variable for AMI vs. stroke on the left panel, and ASCVD death vs. AMI or stroke on the right. Enrichment is calculated as described in the figure caption.

**Figure 7:**
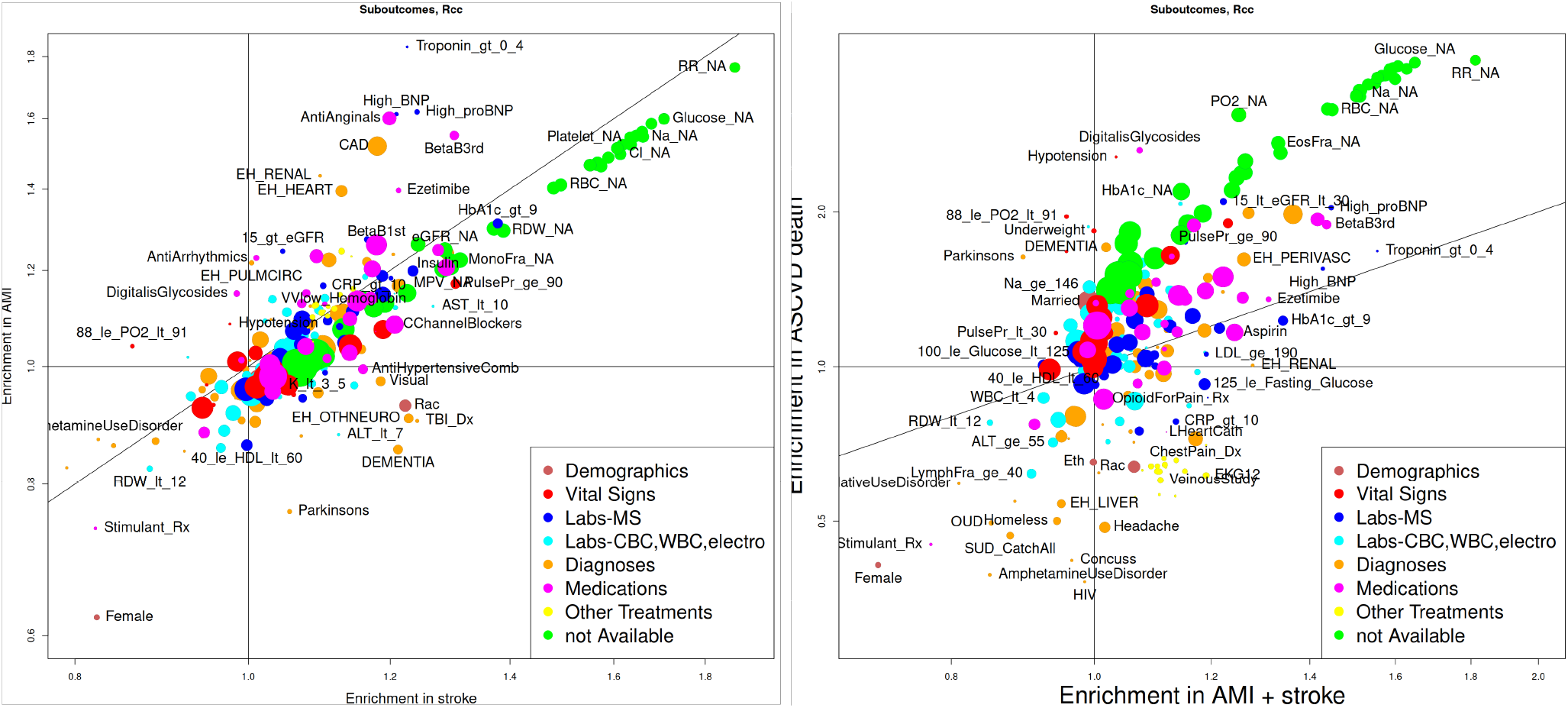
Association of each predictor variable with (left) AMI on the y-axis and stroke on the x-axis, and (right) ASCVD death on the y-axis and AMI + stroke on the x-axis, for the Rcc study design. The color code and symbol sizes are the same as in Figure 6. Note that these are not model coefficients, but the fraction of patients with the indicated suboutcome divided by the number of controls, for each variable, compared to that quantity for all patients. Vertical and horizontal axes and a line of slope one through the origin are provided for reference.

Most variables lie along the diagonal, and are equally enriched in both outcomes. Most clear among them are the large number of laboratory and vital sign metrics not being recorded (green symbols) that are 40% enriched in both AMI and stroke, and 3-fold enriched in ASCVD deaths, compared to the average patient. It is useful to compare this to the coefficients, where these missing variables are protective. It must be noted that patients missing one lab are often missing others, and so this discrepancy between uni-variate associations and model coefficients should be expected. It means the model is using the profile of missing information (labs, vitals, and medications) to implicitly model types of healthcare utilization.

Variables plotted above the diagonal in the left panel are enriched in AMI sub-outcomes, and include high troponin and BNP levels, diagnoses of CAD, heart failure, renal failure, and prescriptions for antianginals, beta blockers, and antiarrhythmics. In contrast, the symbols below the diagonal are enriched in stroke, and include a diagnosis of traumatic bran injury, female gender, Black race, low potassium, HDL between 40 and 60 mg/dl, and diagnoses with dementia or Parkinsons.

The Figure 7 right panel is more complex, and is dominated by the differing levels of enrichment for the different modalities of data (different colored symbols) in ASCVD death compared to AMI or stroke. Missing labs are enriched in ASCVD death, while specific treatments, such as venous studies or EKGs (the tight cluster of yellow symbols at coordinates 1.2,0.6) are more enriched in AMI and stroke. Labs, diagnoses, and medications are generally in lines of slope one, at different levels of enrichment in ASCVD death, with medications most enriched and specific lab results least. This result is concordant with expectation if one assumes an important, confounding, reporting bias that the Dx-code based outcomes (AMI and stroke) are more likely to be identified in patients with a greater degree of VA care, while ASCVD death, which is reported through the CDC’s National Death Index, is independent of VA utilization except for a matching to patient registration records.

### Dependence on confounders

Figure 8 compares the behavior for each component of our MACE outcome to that of selected variables as a function of age (left) and a metric of healthcare utilization (right). While they are not the only confounding variables in the analyses, they do represent the two quantities that were matched in the Rcc study, but not in C-17. Examination of these cases will provide insight into the complexity involved in interpreting the large number of predictor variable.

**Figure 8:**
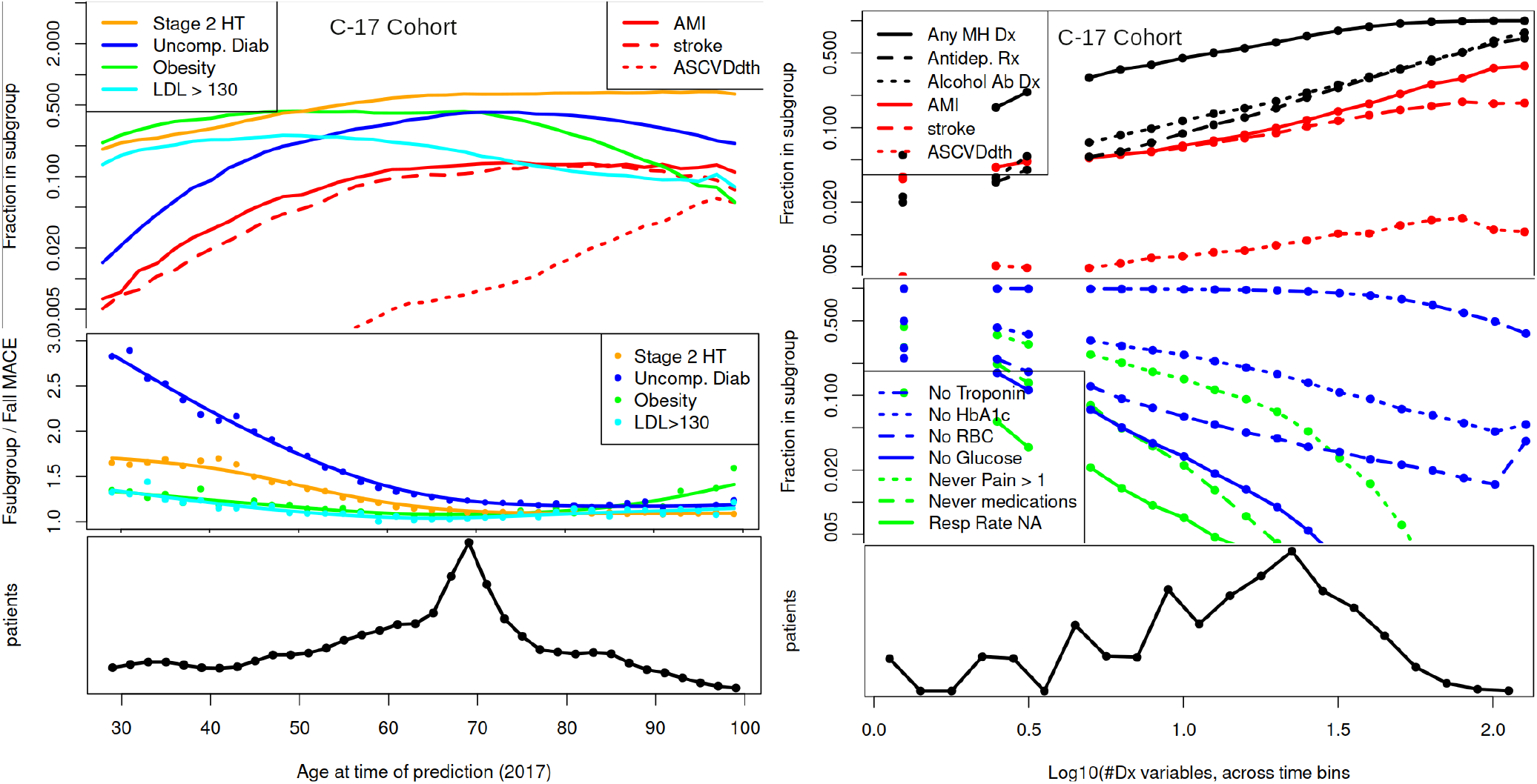
Characterization of two confounding variables for the C-17 cohort: age (left) and an aggregated measure of healthcare utilization (right). The top left panel shows on a logarithmic scale the age dependence of the fraction of each component of our combined outcome (in red), and variables representative of each of four risk factor groups, hypertension, diabetes, obesity, and high LDL cholesterol. The middle left panel compares the age dependent relative risk for MACE for each of the above subgroups vs. the overall C-17 cohort. The bottom left panel is a histogram of total number of patients in each age group. The top right panel shows the fraction of each indicated subgroup on a logarithmic scale as a function of an aggregated measure of healthcare utilization for each component of our combined outcome (in red) and for three behavioral healthcare usage metrics (in black). The middle right panel is similar, but shows the number of patients **not** receiving the indicated laboratory, vital sign, and pharmacy usage. The bottom right panel shows a histogram of the number of patients at each level of healthcare utilization. Our healthcare utilization metric (x-axis, right panel) was computed by summing the number of diagnosis code present over the 47 distinct diagnosis variables across each of the 8 time windows of data collection, for a maximum possible score of 376 (each bin is encoded as presence / absence, or 0/1). We placed the x-axis on a log10 scale, for which the histogram peaks at 1.3, or 20 codes present.

The three components of our outcome are shown as fractions of each age group from 30 to 100 years old, in red in the top left panel. Both AMI and stroke increase exponentially from 0.5% to nearly 10% of the patients as their age at time of prediction increases from 30 to 60 years of age. Above age 60, the fraction of patients having an event in the next five years stays nearly constant at 10%. In contrast, the ASCVD death steadily increases from 0.5% at age 60 to 5% by age 100. As described above, the relevant difference is that AMI and stroke are both reported as diagnosis codes in the VA medical record, while ASCVD death is obtained through the information on death certificates, reported through the CDC’s National Death Index, and then matched to identities of VA patients. As such, it may reflect the impact of reporting differences on outcome determination.

Stage 2 hypertension, defined by a median systolic blood pressure above 140 mmHg in a time window, increases from 20% at age 30 to 50% by age 60. Obesity, defined as a median BMI ≥ 30 kg/m^2^ during a time window, increases to a peak of nearly 50% at age 50, then starting at age 70 the prevalence decreases steadily to about 7% by age 100. Uncomplicated diabetes, defined as a diagnosis code, increases from 1.5% at age 30 to a peak of 45% at just over age 70, then decreases in prevalence. High LDL cholesterol (> 130 *mg/dL*) peaks at age 47, then decreases above that, presumably because of increased statin usage.

The age-dependence of the relative risk four these four factors can be seen in the middle panel, where we present the risk ratio for MACE (defined as the MACE prevalence within each subgroup to the MACE prevalence across all patients) in the C-17 cohort. The strongest relative risk is in the younger diabetic patients, with those under 45 years of age experiencing more than 2-fold elevated MACE risk. With stage 2 hypertension in those under 45, the risk is 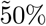 greater compared to those without this diagnosis, while obesity and high cholesterol each confer approximately 20% greater risk. The increased risk associated with these risk factors, other than diabetes, is further augmented above age 70, with obesity showing a strong increase above age 90.

The right panel in Figure 8 compares the incidence of the three components of our MACE outcome to a measure of healthcare utilization, as described in the caption. The incidence of AMI and stroke increases in proportion to the number of recorded variables. This could be due to either a greater amount of healthcare associated with MACE patients or a greater likelihood of identifying people who use the VA healthcare system regularly. This effect can be disambiguated. First, we see that among patients with more than 30 positive variables (1.5 on the x-axis) there are approximately 2-fold more heart attacks than strokes, although both are defined by Dx codes. Second, the NDI-associated ASCVD deaths (not including patients with a recorded AMI or stroke) also increases with increasing healthcare utilization, although with a smaller slope.

In black are shown the incidence of any mental health diagnosis, alcohol use disorder, and the prescription of antidepressants. All have a similar slope to AMI, except that the curve for any mental health diagnosis saturates at about 50 Dx codes. The middle panel shows healthcare utilization in terms of lack of laboratory results, vital signs, and the prescription of any drug. Our models are able to use this set of variables to correct for healthcare utilization patterns in a complex manner. The bottom panel shows a histogram, which is a bumpy because of the binning noise of low counts on the log scale, but otherwise well behaved, with good signal all the way across.

## Discussion

The pathophysiology of CVD progresses throughout a person’s lifetime and the prevalence of clinical CVD has greatly increased in recent years, now accounting for 32% of deaths worldwide [29]. We have attempted to improve CVD prediction in this work by assembling high dimensional longitudinal data sets with carefully encoded predictor variables and millions of veterans in four cohorts with distinct study designs. Our analyses then predicted MACE outcomes with both linear and non-linear models. Our analyses provided statistical power to constrain 243 longitudinal and four demographic variables with any of our prediction methodologies. As with recent work by Zhou [82] and Steele [73], our high dimensional models outperformed our implementation of an AHA-PCE-like model in both discrimination and calibration. Attempts to compare accuracy metrics across published studies, however, are complicated because each study typically makes distinct choices in study populations, choices in variables, their curation, and their encoding, exclusion criteria, and missingness of information [67].

In this paper, we systematically consider our predictor variables, results for measures of disease severity, confounding variables of age and healthcare utilization, co-morbidities, and indicators of patient behavior. We finally assess the effectiveness of interventions, both in terms of included variables and analysis methodologies.

### Predictor variables

We included several modalities of data: laboratory results and vital signs, diagnoses, treatments, and demographics. These domains of data include markers of CVD severity, age, demographics, and patterns of healthcare utilization. For laboratory results, diagnosis codes, and treatments, we included most CVD-specific measures as well as a variety of other commonly used codes. Using the language of Ref. [62], we modeled a wide range of covariates and now compare results to studies that exclude, randomize, stratify, or match on most of the conditions. This allows us to compare subsets of our model coefficients to specific studies exploring a subset of variables. Our linear coefficients, however, are extrapolated from a baseline of real-world levels of comorbidities and treatments, as much as the VA population reflects real-world conditions.

Although we used LASSO model selection [76] as an objective way to control the dimensionality of our fit, our goal was not simply to develop a parsimonious predictive model or an exploratory search for novel predictors as in Ref. [1], but to understand the relative importance of a large number of interpretable variables. Known CVD risk factors include both behaviors (tobacco use, diet, and physical inactivity) and metabolic measures (lipid levels, hypertension, obesity, and diabetes mellitus) [29]. The importance of these risk factors has been established in numerous studies, such as those containing current treatment guidance [33]. Unfortunately, it is difficult to isolate these risk factors from other elements of study design, such as missingness of data, confounding variables, and composition of the study population [9]. Additional challenges arise in CVD prediction because the pathophysiology of the several important metabolic risk factors are intertwined with one another and have been labeled ‘metabolic syndrome’, with origins in obesity, inflammation, and insulin resistance [21]. In this situation, not only it is important to carefully define a study design with appropriate predictor variables [14], but also to recognize that missing interaction terms can introduce spurious relationships in linear models [71].

To facilitate the comparison of model coefficients across data types, we encoded all variables except age as binary (zero-one) variables. For laboratory results, it is helpful to define a spanning, non-overlapping set of categories, including NA as a distinct category. We did this using standard ranges for low, normal, and above normal categories. Collins *et al*. [8] showed accuracy improvements of only a few patients per thousand for continuous variables over 3 or 4 categories, so we chose the improved interpretability, decreased sensitivity to outliers, and ability to relate directly to terms used in standards of care. ICD billing codes are used by the VA to capture patient diagnoses, and the potential problems in directly using them to characterize a state of health have been outlined [75] and assessed [57]. The changeover in the US from ICD9 to ICD10 systems on October 1, 2015 further complicates their use [49]. To account for these difficulties, we utilized an extensive set of derived Dx-based variables that had already been harmonized across the ICD9/ICD10 changeover, from McCarthy, et al. [52], and updated them according to our own analyses needs. For medications, we simply grouped medications according to the VA Drug Classification System.

Another important aspect of variable definition in time-dependent studies is the treatment of time periods and missing data. Laboratory results and vital signs (except pain) were treated as observations, and carried forward, then backwards, across the time windows of data collection for a given patient. All diagnosis-based, procedure, and medication data were treated as events, in that absence of a record was treated as absence of the variable. Clearly, this is more appropriate for, e.g., overdoses or an insulin fill than for a diagnosis of uncomplicated diabetes.

### Risk profiles

Our curation and harmonization of the laboratory results enables us to understand the relative likelihood of MACE events for different levels of a range of laboratory results, and to compare the importance of abnormal labs and vital signs across a breadth of risk factors. Our Rcc study design matches on birth year and level of healthcare utilization, and we also control for other healthcare utilization patterns, some behaviors and varying levels of treatment. It is reasonable to compare our Rcc models to previous outcomes analysis to constrain potential pathophysiological mechanism for each of the lab/vital sign based predictors, and we begin with that process across the range of risk factors. Many risk factors are independent predictors of comparable amplitude, and we grouped them in Figure 4 for the metabolic variables and in Figure 5 for variables relating to disease progression and mental health conditions.

#### Lipid metabolis

includes measures of HDL cholesterol, LDL cholesterol, total cholesterol, and triglycerides, and is reviewed in the context of CVD in Reference [72]. Elevated LDL cholesterol is a major cause of heart disease, causing both atherosclerosis and inflammation. We observe a steady increase of MACE risk with greater LDL concentrations, and even a slight decrease in risk from below-normal LDL levels. HDL particles promote cholesterol efflux and reduce inflammation. HDL metabolism is more complex, with lower MACE risk associated with high HDL levels, as long as both triglycerides and LDL cholesterol are not elevated. As seen in Figure 4, very high levels of HDL cholesterol are predictive of MACE events, as reported by others [47]. Two studies carefully examined the probability of MACE outcomes on lipid levels [48, 47] and comparison of their Figure 1 to our coefficients characterizing lipid dependence shows striking agreement on the non-linear (U-shaped) effect of HDL and LDL on MACE outcomes. As we found, elevated triglycerides were also linked to elevated atherosclerotic cardiovascular disease risk [34, 72].

#### Hypertension

has long been known to be among the important CVD risk factors, with the stresses on the arteries causing a similar type of damage to atherosclerosis, but in a complementary manner [37]. It is a stronger risk factor in younger adults and the risk rises steeply for those with a systolic blood pressure of >140 mmHg (stage 2 hypertension) [42], as we saw in Figures 4 and 8. While systolic blood pressure continues to increase with age, diastolic tends to decrease, and our observation of high pulse pressure being an independent MACE predictor is consistent with the literature [18]. We found that hypotension, although present only in a small minority of patients, was also a strong predictor of MACE events. This observation has been controversial for some time, especially in terms of aggressiveness of therapy to reduce high blood pressure [50, 79]. A meta-analysis by Bohm *et al* [6] showed that a target blood pressure of 120–130 mm Hg systolic and 70–80 mm Hg diastolic is associated with the lowest rate of CVD events. We also observed a high pulse/heart rate to be independently predictive of MACE events, in agreement with Ref. [74], although we did not observe elevated risk for low heart rates.

#### Decreased renal function

measured by reduced eGFR and increased urinary albumin-to-creatinine ratio were shown to be independent predictors of MACE events in type-2 diabetics in the ADVANCE study with a 2.2 hazard ratio for cardiovascular events for every halving of eGFR [56]. As seen in Figure 4, reduced eGFR is an acute MACE predictor and MACE risk increases at eGFR levels of 60 and below. Although our coefficients in Figure 4 were small (0.1-0.2), 44% of the patients in Rcc with an eGFR between 45 and 60 also had a diagnosis of CAD, with a model coefficient of 1.0. A follow-up retrospective study by Currie, *et al*. [15] reported that the MACE risk for a given level of eGFR was 2.4-4.6 times higher in the presence of type-2 diabetes. The importance of understanding the interdependencies of risk factors is evident in meta-analyses of cardiovascular outcome trials with SGLT-2 inhibitors to reduce CVD events and mortality [31, 32]. Understanding these is even more important in type-2 diabetes patients with impaired renal function, where reductions ranging from 7% to 20% beyond standard of care were achieved, but the relevant differences among trials was still unclear [31, 32].

#### Electrolyte imbalance

are present in only a few percent of the participants in Rcc, but their model coefficients are as large as the other predictors we have discussed, with low sodium, potassium, magnesium, and chloride predictive of MACE events and low calcium levels protective. Low sodium [64] and potassium and magnesium [43] levels are associated with poor outcomes in CVD patients. Community prevalence and the distinct risk factors of the various electrolyte disorders are discussed in Ref. [45]. Numerous classes of medications are available to treat hypertension, and electrolyte imbalance is one of several important side effects considered in the choice of medication [36]. The protective effects of low serum calcium levels with respect to MACE outcomes that we observe may be related to the direct appearance of Ca^2+^ ions in the aterial plaques, promoting both stroke [80] and AMI [38].

#### Diabetes

has been reported to increase women’s chronic heart disease risk by a factor of approximately 2.8, and men’s by 2.2 [60]. Although the presence of diabetes is highly correlated with many other predictor variables, mechanistically, it augments microvascular and macrovascular risk and is considered to be an independent predictor of many vascular disease outcomes [44]. The risk coefficients in Figure 4 show that both high HbA1c and high glucose are predictive of MACE events, and that the effect is positively associated with blood glucose levels. Only rarely is hypoglycemia seen, but when it occurs it is strongly predictive of MACE events. Both diabetes diagnostic codes and medications protect against MACE events, and our interpretation is that this is because diagnosis and treatment are highly correlated. Our results are consistent with the observation of Galbete *et al*., who noted in a review of 19 studies of CVD risk in populations with diabetes and 46 studies of the general population that many studies have a high risk of bias due to methodological shortcomings and scarcity of independent validations [28].

#### Obesit

in our study is protective against MACE events (Figure 4), despite the general epidemiological association of obesity with an increased risk of developing CVD [25]. The observation that being overweight or obese can improve chances of survival is known as the obesity paradox [7]. While the association of obesity with the metabolic syndrome [21] and its co-linearity with other MACE risk factors certainly reduces our observed model coefficients, this effect is still observed in an univariate analysis and across multiple study designs. Several effects appear relevant to this point. First, we observe, in agreement with Ref. [59], that being underweight (BMI *<* 18.5 *kg/m*^2^) is a risk factor for CVD, perhaps due to poor nutritional status. Second, BMI as a predictive variable, ignores important issues of muscle mass and the distribution of adipose tissue that strongly impacts the prognosis associated with obesity [25]. Finally, selection biases are likely occurring because we restricted our analysis to the first recorded MACE event. The observation that obesity has a much larger model coefficient in Rcc than C-17 supports this supposition.

#### CVD progression

is not considered in the AHA-PCE equations [33], presumably because of their co-linearity with other risk factors and the difficulty in defining an appropriate cohort in a generalizable manner. Nevertheless, our observed coefficients are in line with those for CAD [54] and PVD [3] of 1.3-3.0.

#### Other aspects of pathophysiology

have been explored in our model, such as nicotine dependence, delivery of estrogen, complete blood counts, and differential blood counts. Most of them have smaller model coefficients and lower prevalence, compared to the major risk factors described above, but can themselves be related to the literature in a more detailed treatment of this data set.

Our comparison of risk factors for **Stroke vs. AMI** agreed with Reference [30] that hypertension and being female were more favored in stroke, but also identified Blacks, Hispanics, and pulse pressure > 90 mmHg, visual impairment, traumatic brain injury, dementia, and Parkinson’s as more associated with stroke than AMI. Diagnoses of CAD, chest pains, renal failure, and heart failure, taking anti-anginals and 3rd generation beta blockers, and laboratory results with elevated troponin were more associated with AMI. We did not see the association of cholesterol and diabetes with AMI that was observed by [30].

#### Age

is a strong predictor of both MACE events and its four main risk factors, as shown in Figure 8. Additionally, essentially all of our predictor variables and many competing causes of death that we did not include are associated with age to some degree. As a straightforward consequence, model coefficients of predictor variables will be dependent on how age is treated in the model, and the predictive ability of a MACE model may change significantly with the age distribution of the model cohort or subgroup being examined. We explore this association in sensitivity analyses by including a date-of-birth cohort matched study design, Rcc, in our analyses along with the three non-matched study designs. Figure S1 shows the explicit age-dependence observed differs considerably between models, while Figure 3 shows excellent calibration in subgroups for both Rcc and C17, demonstrating both models can predict MACE events well.

The situation is not symmetric, however. Figure 2 shows that in a GLM model trained on Rcc the c-statistic slips from 0.68 to 0.62 when evaluation shifts from Rcc to C17. Similarly, in a GLM trained on C-17 the c-statistic falls from 0.75 to 0.55. Although some of this degradation in discrimination is due to the different underlying risk distributions, as described in Ref. [11], it is evident that the Rcc-trained model generalizes better to C17 than the reverse. Figure 6 shows that while the two study designs differ across many model coefficients, the largest differences involve metrics of healthcare utilization, such as not having respiratory rate measured, or never reporting pain. This result prompted us to examine how much model accuracy could be recovered by re-optimizing the tradeoff between age and healthcare utilization patterns, described in detail in the caption to Table S1. Indeed, much of the model accuracy can be recovered in this way, and reoptimized Rcc-trained models produce discrimination scores on C17 within two percentage points of C17 trained models.

#### Healthcare utilization patterns

introduce confounding effects into our study as 90% of our outcomes and nearly all of our predictor variables are extracted from the EMR. Unlike age, healthcare utilization can be assessed by a variety of metrics, such as visits, medications, common laboratory results, specialty labs, or utilization of preventative medicine. We elected to use minimal exclusion criteria to enable the model to identify the best markers of utilization, and many variables emerge as important predictors of MACE outcome (see Figure 8). One interesting aspect of the non-linear models is that they are capable of allowing model coefficients to depend on the level of available information. Thus, while the LDL-DS trained model (where LDL cholesterol labs are an inclusion criteria) outperforms Visit-DS (where only a visit is required) by 3 percentage points, little difference is observed among the non-linear models.

Further understanding of healthcare utilization biases can be found by comparing predictors of our sub-outcomes: specifically ASCVD death (which is reported independently of the EMRs) to AMI and stroke (which are obtained from the EMRs). The propensity of different modalities of data to be associated with ASCVD death rather than Dx-encoded outcomes is strongest for missingness of laboratory results and weakest for CVD-specific procedures, with other modalities of data intermediate in the spectrum, visible in Figure 8. Nevertheless, Figure 8 shows that even the likelihood of ASCVD death increases in proportion to greater levels of healthcare utilization. Our interpretation of this finding is that the level of healthcare utilization predicts MACE outcomes both because of reporting biases and because patients with CVD have a greater level of healthcare utilization.

### Predictive models

Model performance metrics such as c-statistic are often used to compare models. We have seen in Figure 2 that different prediction methodologies provided the highest score in the four different cohorts. Evidently, the best methodology is dependent on choices in study design, completeness of data, included predictive variables, confounding variables and definition of outcome. Alternative metrics to assess model performance could include subgroup analysis, calibration, and comparison of model coefficients with known mechanisms. We found that the three non-linear models typically performed better than logistic regression with subgroup analysis and calibration. Linear model coefficients characterizing laboratory results and vital signs were generally consistent with literature, across all study designs.

Other variables such as age and healthcare utilization depend strongly on cohort and study design, raising concerns for both the generalizability of the model and the interpretation of model coefficients, as discussed in the literature [78]. This observation motivated our Rcc study design, which matches on two of the strongest confounding variables. While techniques exist to compare predictor variables to mechanisms for non-linear models [66], these methods themselves are complex and we chose not to explore them further here.

Model generalizability, broadly defined in [39], is a valuable performance metric for models, but done properly would require repetition of this entire work in another healthcare system. By comparing across study cohorts, and the transferability of models across cohorts, with re-optimization, we gain understanding of which models will generalize better and why. The Rcc-trained model outperformed the C-17-trained model in two respects. First, it retained much more of its accuracy when transferred across cohorts, both before and after re-optimization. Second, the improvement in model scores with the nonlinear models (NN, tabnet, RF) over logistic regression was twice as large for Rcc compared to the other study designs. Our Rcc study design matches on two of the strongest confounding variables, and forces the nonlinear models to evaluate interaction terms among more interesting variables, such as comorbidities and between modalities of data, such as diagnoses and treatments. One important aspect of the problem we did not explore is accounting for the possibility of repeated MACE events.

If the internal representation of the data is an important determinant of a model’s utility, at least two basic strategies can be found in the literature. Pham *et al*. [61] provides an elaborate scheme to reproduce the logic of disentangling outcomes into component concepts, utilizing many of the sophisticated forms developed with neural network techniques. Without proper levels of data completeness and ability to control for confounding and biases, however, it is unclear whether thee concepts will decompose according to the intended logical decomposition. Another approach is to explicitly model the joint probability distribution of two explicit variables. Such joint models have been used to model treatment effects [63], repeated intermediate outcomes [58], or correlated outcomes [24]. Clearly, there are many possibilities for how to set up such models, and the ‘best’ solution will likely depend on the problem in question, the available data, and the intended use of the model.

## Conclusions

We have compared high-dimensional predictions of MACE incidence in millions of US Veterans across study designs and prediction methodologies. Our calculations provided sufficient statistical power to constrain 243 time-resolved variables representing age and demographics, laboratory results and vital signs, diagnoses and treatments, and several measures of healthcare utilization. Predictors characterizing the pathophysiology of CVD, including obesity, lipid metabolism, diabetes, hypertension, CAD, PVD, and comorbidities remained relatively consistent across study designs and associated with MACE events. Laboratory measures and vital signs provided risk profiles consistent with, but difficult to quantitatively compare to, existing literature. Diagnosis codes related to laboratory results tended to be protective, presumably because they were indicators of patients seeking treatment. Medications were mixed in their association with MACE outcomes, with the presence of any medication being strongly protective, insulin or high intensity statins associated with poorer outcomes, and metformin, lower intensity statins, and alpha blockers associated with reduced levels of MACE outcome.

The confounding variables of age and those related to healthcare utilization had large coefficients, but these coefficients changed with study design, particularly between Rcc (which was year-of-birth matched) and C-17 (which included all patients active in a 3-month period). This change suggests that associations with homelessness, alcohol dependency, obesity, non-zero pain, not having respiration rate measured, and not taking medications may be due more to selection biases than their direct impact on MACE risk.

Comparison of model discrimination and calibration across a dozen subgroups showed significant improvement for all high-dimensional models in comparison to our baseline AHA-PCE-like model, and greater performance in the non-linear models compared to GLM models. The Rcc study design, enriched in cases, and with prediction time set to six weeks before the predicted event, the LDL-DS (Cox) model, and the C-17 model with 5-year interval prediction, all provided accurate calibration and comparable discrimination, although with different methods of encoding the longitudinal aspect of prediction. The LDL-DS and Visit-DS study designs, with 12 years of follow-up, did not perform well with interval prediction methods, and the LDL-DS model, which was trained on fewer patients, but with more complete laboratory data, outperformed Visit-DS, even on the Visit-DS study design. Models trained on Rcc could be re-optimized using age and a handful of other variables, including healthcare utilization, to achieve performance on the C-17 cohort comparable to models trained on C-17, while the reverse process was much less effective. The ability of re-optimized Rcc-trained non-linear models to approach the level of discrimination of C-17 trained models on the C-17 cohort suggests a greater generalizability for models trained in this manner. Our neural network model outperformed our GLM by four percentage points in Rcc, which had acute information for all events.

By treating the wide range of risk factors in a single calculation, we demonstrate a broad set of independent predictors for the risk of MACE outcomes, designed to be interpreted in the presence of the other factors. Comparison across study designs and prediction methodologies highlights variables whose impact on CVD risk is less likely to generalize from the present study population or the VA healthcare system, such as age and measures of healthcare utilization. While it would be valuable to clearly distinguish the effects of laboratory results, diagnoses, and treatments, we were not able to achieve this level of model resolution in this work; more sophisticated longitudinal study designs with distinct models of within-patient and between-patient parameter variation will be required to solve this important class of problems.

## Data Availability

All calculations underlying figures will be made available in SI. Underlying EMR contain PHI and cannot be made available.

## Acknowledgements

We would like to thank the many people involved in setting up the MVP-Champion infrastructure at Oak Ridge National Laboratory, including Sumitra Muralidahar, Dimitri Kusnezov, Edmon Begoli, and Franciel Linarus and team. We would also like to thank Jodie Trafton and her team at the VA Office of Mental Health and Suicide Prevention for providing ICD-9 and ICD-10 codes for their 100 Dx-related variables and much other pragmatic modeling advice. We would like to thank numerous VA and DOE colleagues who developed these ideas over the past several years, and Mac Hyman for carefully reading many previous iterations of this manuscript. This research was conducted with the approval of the VA Central IRB with project number MVP014. This research is based on data from the Veterans Health Administration and was supported by the MVP-CHAMPION collaboration between the Department of Energy and Department of Veterans Affairs. This publication does not represent the views of the Department of Veteran Affairs, Department of Energy, or the United States Government.

## 1 Supplementary Information

**Table S1:**
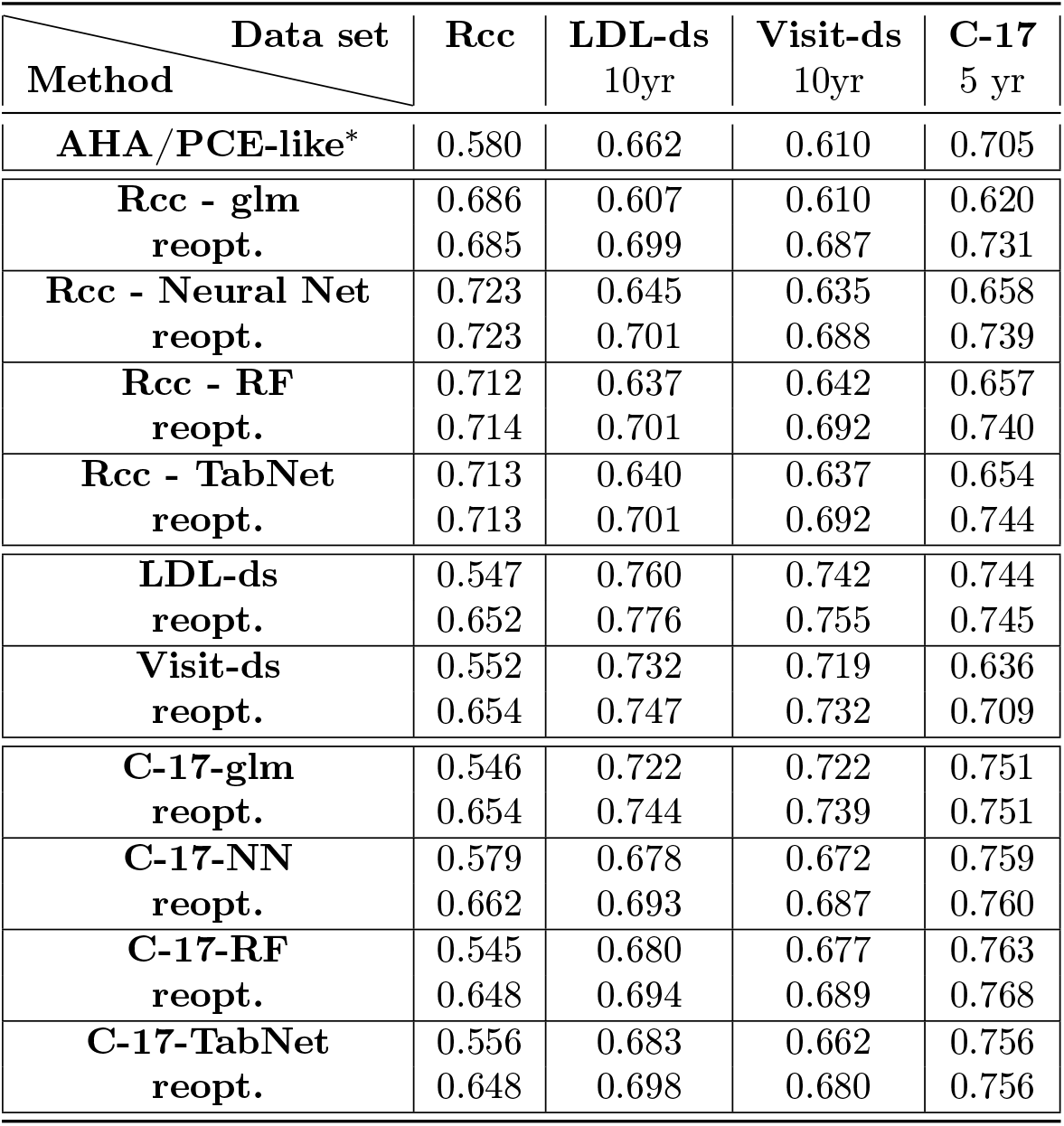
C-statistics for MACE prediction from a variety of study designs and prediction methodologies, including an assessment of the transferability of models from one study design to another. The first row provides C-statistics for an AHA-PCE-like model for the indicated study designs, where models were trained on half the data and evaluated on the other half for each study design. For Rcc and C-17, models were trained with four prediction methodologies (GLM, NN, RF, TabNet), and evaluated in each of the study designs. Within each table cell, the top number indicates C-statistic with the trained model used directly on each study design, while the lower number is the C-statistic with each model score included together with Gender, Race, Ethnicity, Age, Age^2^, Pain gt 0, AnyDrug, HbA1c gt 9%, DEMENTIA, EH HYPERTENSION in a glm re-optimization. Typically, this re-optimization improves the score by less than two percentage points, but for the case of Rcc models used to predict on the non-age-matched models, model performance increases by 8-10 percentage points. About half the improvement comes from the age variable. For our time-to-event models (LDL-DS and Visit-DS), scores were evaluated as interval predictions at 10 years past time of prediction.

**Table S2:** Attributes of selected predictor variables. For each of 40 variables shown, we provide 11 columns of information, including the percentage of total patients in each category (% tot), logistic regression model coefficients for C-17 (C.C17), Visit-DS (C.V), and Rcc (C.Rcc) cohorts, the calculated odds ratio for Rcc (exp(*C.Rcc*) in OR.Rcc), number of cases and controls in the Rcc cohort, the percentage of cases divided by the percentage of controls for each variable, and an enrichment factor for each element of our combined outcome (Enr.AMI, Enr.Stroke, and Enr.dth), computed as the number with the respective outcome divided by the number of controls for each variable, in turn divided by that quantity for all patients.

| Conditions | % tot | C.C17 | C.V | C.Rcc | OR.Rcc | Cases | Controls | %Cas/%Cnt | Enr.AMI | Enr.S | Enr.dth |
| --- | --- | --- | --- | --- | --- | --- | --- | --- | --- | --- | --- |
| Total |  |  |  |  |  | 1,078,498 | 1,648,330 |  | 31.5% | 26.7% | 7.2% |
| Female | 3.4 | -0.31 | -0.34 | -0.47 | 0.62 | 33102 | 59619 | 0.7 | 0.62 | 0.82 | 0.41 |
| Black | 11.5 | 0.13 | 0 | 0.01 | 1.01 | 151916 | 160427 | 0.97 | 0.93 | 1.22 | 0.64 |
| Hispanic | 3.6 | 0 | -0.07 | -0.07 | 0.93 | 45140 | 51951 | 0.93 | 0.95 | 1.05 | 0.65 |
| Alcohol depen. | 11.4 | 0 | 0.02 | -0.33 | 0.72 | 140322 | 171104 | 0.89 | 0.9 | 1.01 | 0.73 |
| Nicotine Dx | 20.6 | 0.47 | 0.41 | 0.38 | 1.46 | 285800 | 275297 | 1.03 | 1.11 | 1.12 | 0.97 |
| OBESITY Dx | 23.3 | -0.08 | -0.13 | -0.37 | 0.69 | 291662 | 343753 | 0.88 | 0.98 | 0.95 | 0.8 |
| CAD Dx | 23.8 | 0.87 | 0.58 | 1.03 | 2.79 | 393600 | 254971 | 1.35 | 1.52 | 1.18 | 1.98 |
| ChestPain Dx | 15.9 | 0.47 | 0.16 | 0.59 | 1.81 | 226909 | 205638 | 1.06 | 1.22 | 1.11 | 0.73 |
| Hypertension 2 | 43.3 | 0.53 | 0.33 | 0.43 | 1.53 | 598373 | 581602 | 1.04 | 1.04 | 1.14 | 1.32 |
| PulsePr ≥ 90 mmHg | 6.5 | 0.06 | 0.22 | 0.17 | 1.18 | 100079 | 78377 | 1.14 | 1.17 | 1.31 | 1.9 |
| HYPERTENS Dx | 54.8 | 0.15 | 0.1 | -0.02 | 0.98 | 749502 | 745751 | 1.01 | 1.03 | 1.1 | 1.32 |
| Aspirin | 21.4 | 0.16 | 0.26 | 0.27 | 1.31 | 322468 | 262152 | 1.15 | 1.21 | 1.29 | 1.17 |
| ACEInhibitors | 32.8 | 0.07 | 0.07 | 0.11 | 1.12 | 469526 | 423910 | 1.1 | 1.13 | 1.15 | 1.38 |
| AntiAnginals | 11.7 | 0.16 | 0.22 | 0.34 | 1.4 | 197898 | 119921 | 1.31 | 1.6 | 1.2 | 1.93 |
| BetaB 1st gen | 31.2 | 0.19 | 0.17 | 0.27 | 1.31 | 470978 | 380469 | 1.2 | 1.26 | 1.18 | 1.49 |
| Na < 134 | 6.2 | 0.1 | 0.22 | 0.14 | 1.15 | 87795 | 80740 | 1.05 | 1.08 | 1.18 | 1.41 |
| K < 3.5 | 4.6 | 0.15 | 0.13 | 0.23 | 1.25 | 61277 | 63932 | 0.98 | 0.96 | 1.15 | 1.04 |
| Cl ≤ 92 mEq/L | 0.9 | 0.06 | 0.29 | 0.16 | 1.17 | 13449 | 11441 | 1.08 | 1.11 | 1.18 | 2.07 |
| TotChol ≥ 220 mg/dl | 6.5 | 0.17 | 0.12 | 0.21 | 1.24 | 89184 | 86983 | 1.01 | 1.09 | 1.11 | 1.05 |
| LDL 130-160 mg/dl | 9.9 | 0.06 | 0.06 | 0.06 | 1.06 | 128165 | 140553 | 0.95 | 0.98 | 1.05 | 0.98 |
| LDL 160-190 mg/dl | 3.7 | 0.16 | 0.15 | 0.16 | 1.17 | 51600 | 50506 | 1.01 | 1.08 | 1.12 | 1 |
| LDL ≥ 190 mg/dl | 1.4 | 0.19 | 0.22 | 0.24 | 1.27 | 20351 | 17725 | 1.07 | 1.18 | 1.2 | 1.06 |
| Trig ≥ 200 mg/dl | 24.1 | 0.15 | 0.12 | 0.15 | 1.16 | 329803 | 327522 | 1.01 | 1.09 | 1.07 | 1.04 |
| Statin High | 20.8 | 0.02 | 0.06 | 0.14 | 1.15 | 307052 | 260463 | 1.12 | 1.2 | 1.17 | 1.4 |
| Statin Medium | 23.4 | -0.08 | -0.03 | -0.04 | 0.96 | 317539 | 319767 | 0.99 | 1.04 | 1.08 | 1.3 |
| UNCDIAB Dx | 26.9 | 0.02 | 0.01 | -0.15 | 0.86 | 375857 | 357250 | 1.04 | 1.08 | 1.13 | 1.29 |
| COMDIAB Dx | 13.7 | 0.12 | 0.07 | -0.01 | 0.99 | 199280 | 172997 | 1.09 | 1.17 | 1.21 | 1.18 |
| Metformin | 16.4 | -0.02 | -0.02 | -0.06 | 0.94 | 227720 | 218833 | 1.03 | 1.09 | 1.14 | 0.99 |
| Sensitizers | 13.6 | 0 | 0 | -0.06 | 0.94 | 196330 | 175201 | 1.07 | 1.14 | 1.17 | 1.35 |
| Insulin | 9.7 | 0.02 | 0.05 | 0.02 | 1.02 | 148362 | 116546 | 1.14 | 1.25 | 1.28 | 1.36 |
| HbA1c 8-9% | 8.4 | 0.16 | 0.16 | 0.16 | 1.17 | 125231 | 105006 | 1.1 | 1.2 | 1.24 | 1.19 |
| HbA1c ≥ 9% | 6.5 | 0.63 | 0.47 | 0.52 | 1.67 | 102192 | 73862 | 1.18 | 1.31 | 1.38 | 1.23 |
| Glucose > 150 mg/dl | 18.6 | 0.28 | 0.26 | 0.21 | 1.23 | 269424 | 238577 | 1.08 | 1.15 | 1.19 | 1.25 |
| RR NA | 5.7 | 0 | 0 | 0.45 | 1.57 | 112236 | 42472 | 1.51 | 1.76 | 1.87 | 3.94 |
| AnyDrug | 66.5 | 0.04 | -0.07 | -0.88 | 0.41 | 869836 | 942797 | 0.69 | 0.98 | 1.03 | 1.2 |
| EKG12 | 2.7 | 0.27 | 0.22 | 0.47 | 1.6 | 38576 | 34394 | 1.06 | 1.24 | 1.13 | 0.62 |
| Troponin NA | 65.7 | -0.25 | -0.07 | -0.21 | 0.81 | 879744 | 912713 | 0.85 | 0.99 | 1.07 | 1.41 |
| Troponin > 0.4 | 0.3 | -0.08 | -0.01 | 0.07 | 1.07 | 5829 | 2926 | 1.33 | 1.83 | 1.23 | 1.68 |
| WBC ≥ $12 \times 10^9/L$ | 5.1 | 0.13 | 0.21 | 0.16 | 1.18 | 72137 | 67169 | 1.04 | 1.14 | 1.1 | 1.23 |
| EEstradiol | 0.1 | -0.2 | -0.3 | -0.22 | 0.8 | 849 | 2720 | 0.48 | 0.38 | 0.56 | 0.05 |

**Figure S1:**
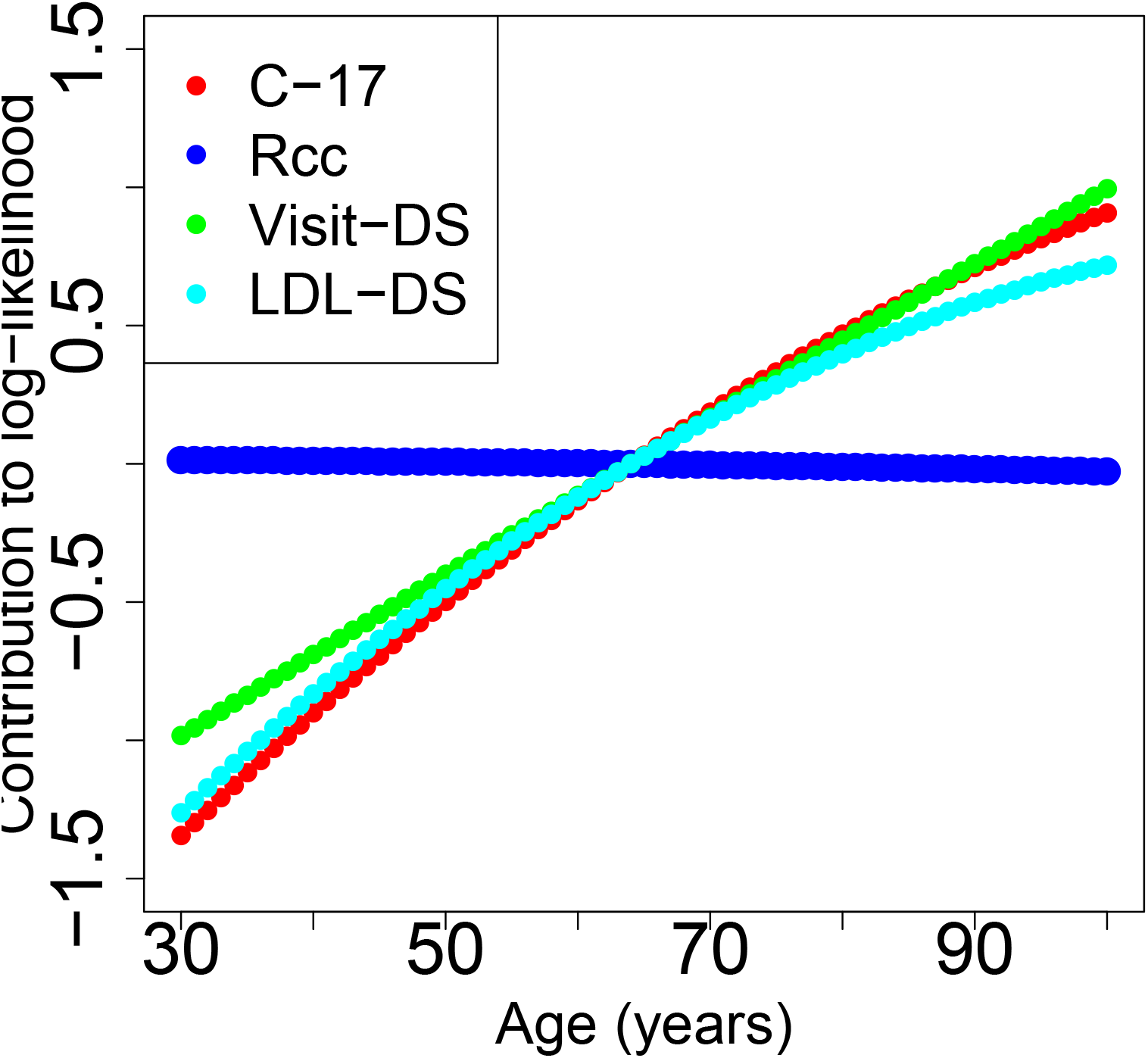
Age dependent contribution to log likelihood of MACE events from age and age^2^ terms of the generalized linear models for each of the four study designs.

**Figure S2:**
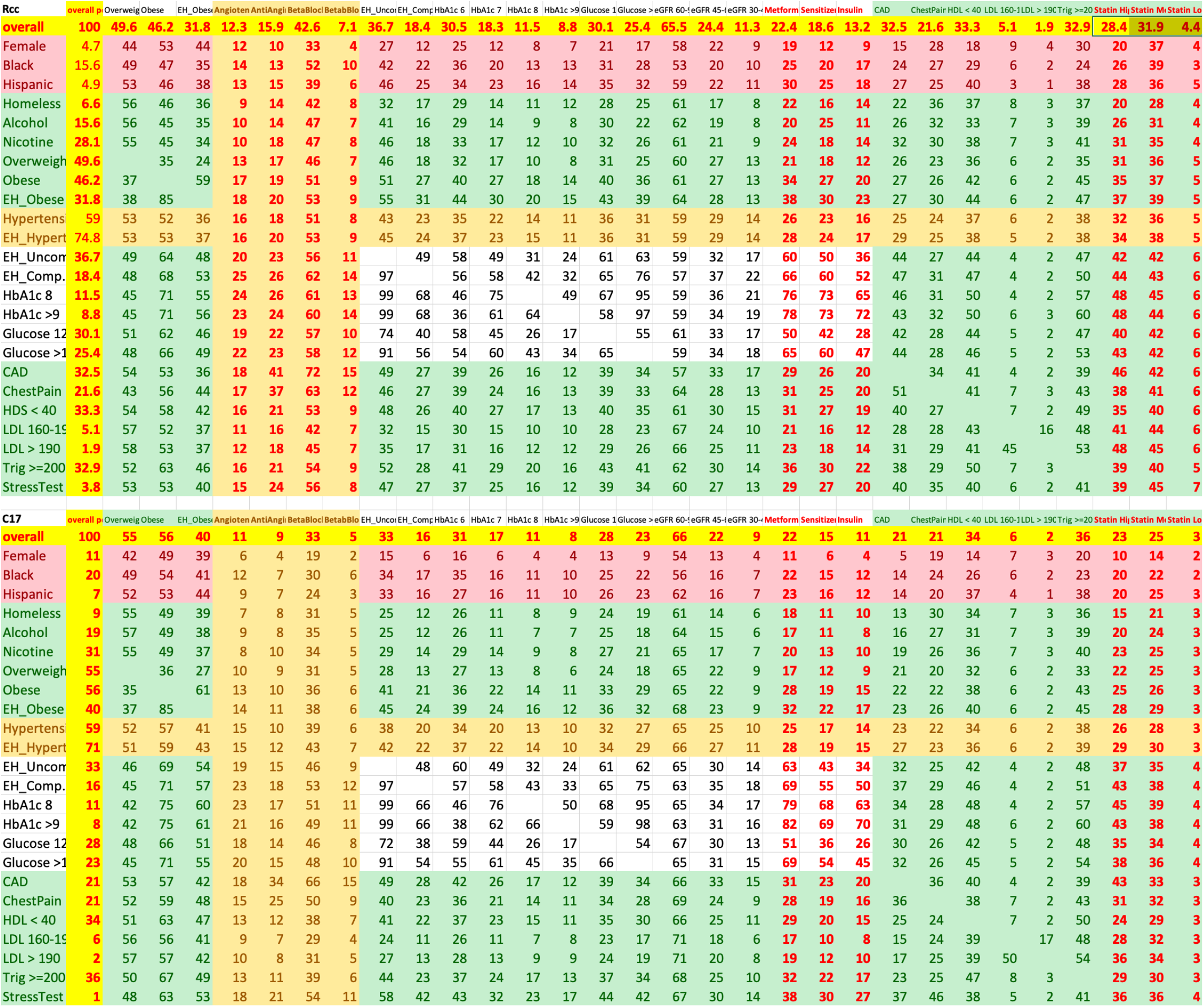
Co-occurrence of selected variables in the Rcc (top) and the C-17 (bottom) cohorts. Each row defines a subgroup of patients, where the value in each column is the percentage of that subgroup with the indicated attribute. For example, 12% of homeless patients have at least one time window with a median HbA1c level above 9%, compared to 8.8% of all patients in the Rcc cohort. Note that Rcc is enriched to 50% cases and is matched on date of birth cohort. With this information we can understand which interaction terms among variables may be important to MACE risk prediction.

**Figure S3:**
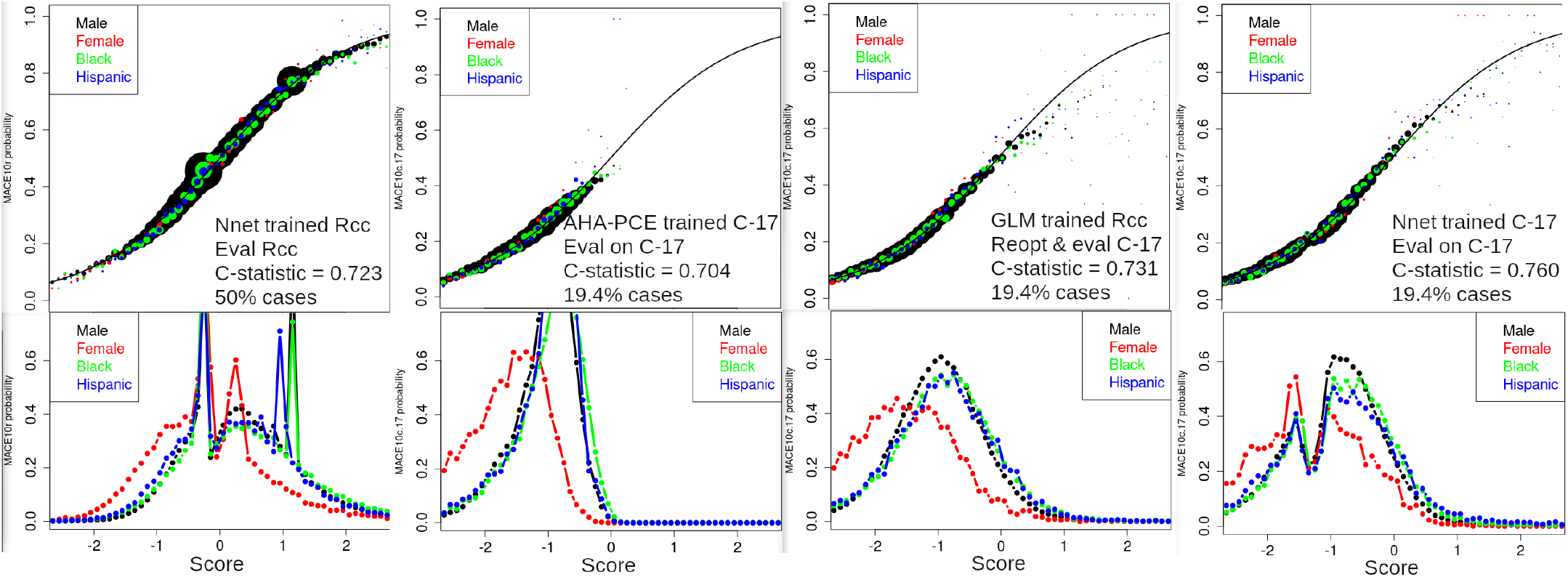
Plots are as described in Figure 3, except with subgroups defined by sex (male and female), race (Black), and ethnicity (Hispanic).

**Figure S4:**
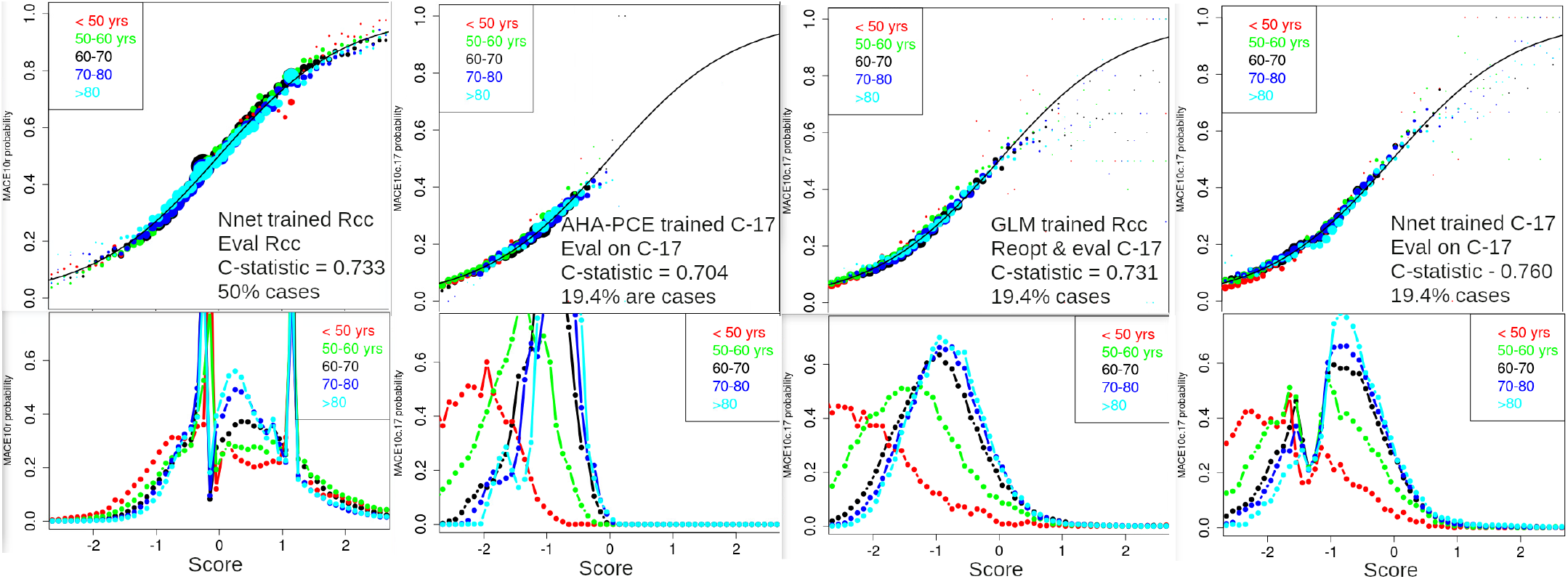
Plots are as described in Figure 3, except with subgroups defined by five age groups, as indicated by color in the legend. Note that the lack of an age dependence for the Rcc study design (left column, bottom panel) is expected, as controls were associated with cases by matching on date of birth.

**Figure S5:**
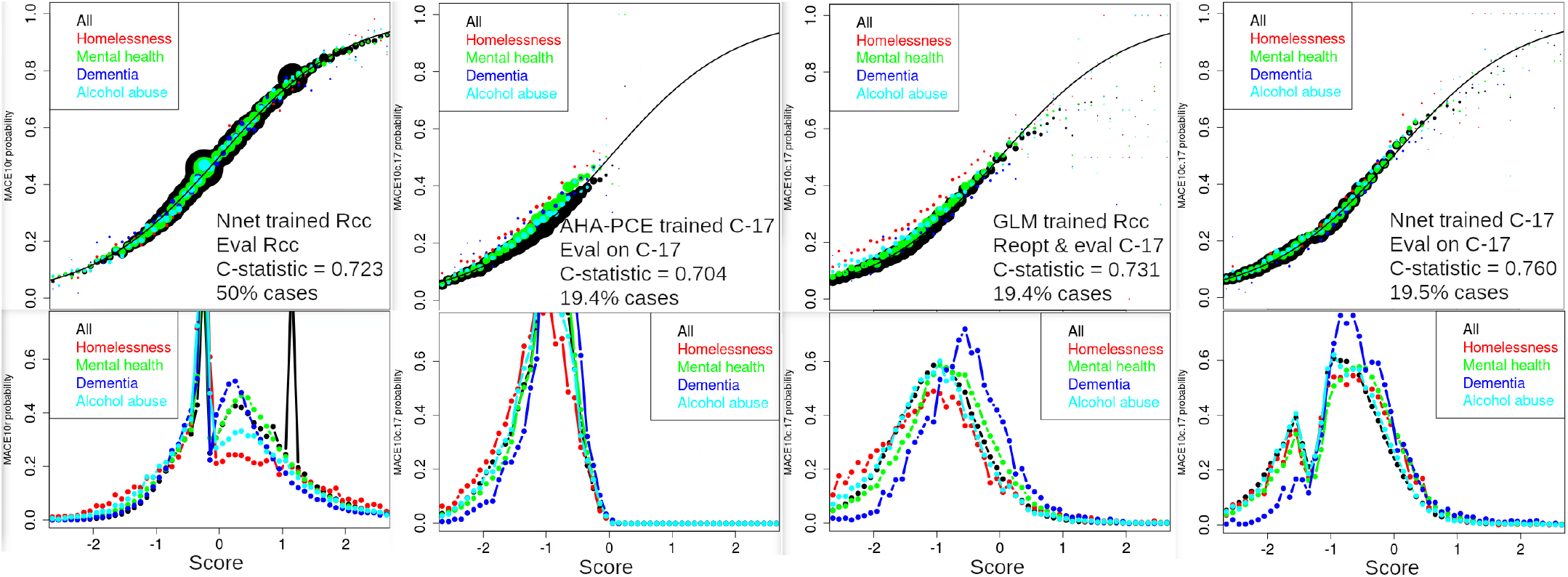
Plots are as described in Figure 3, except with subgroups defined by all patients (black) and four subgroups of patients with potentially distinct healthcare utilization patterns: homeless patients, patients with a mental health diagnosis, patients with dementia, and patients with a diagnosis involving alcohol use disorder.

**Figure S6:**
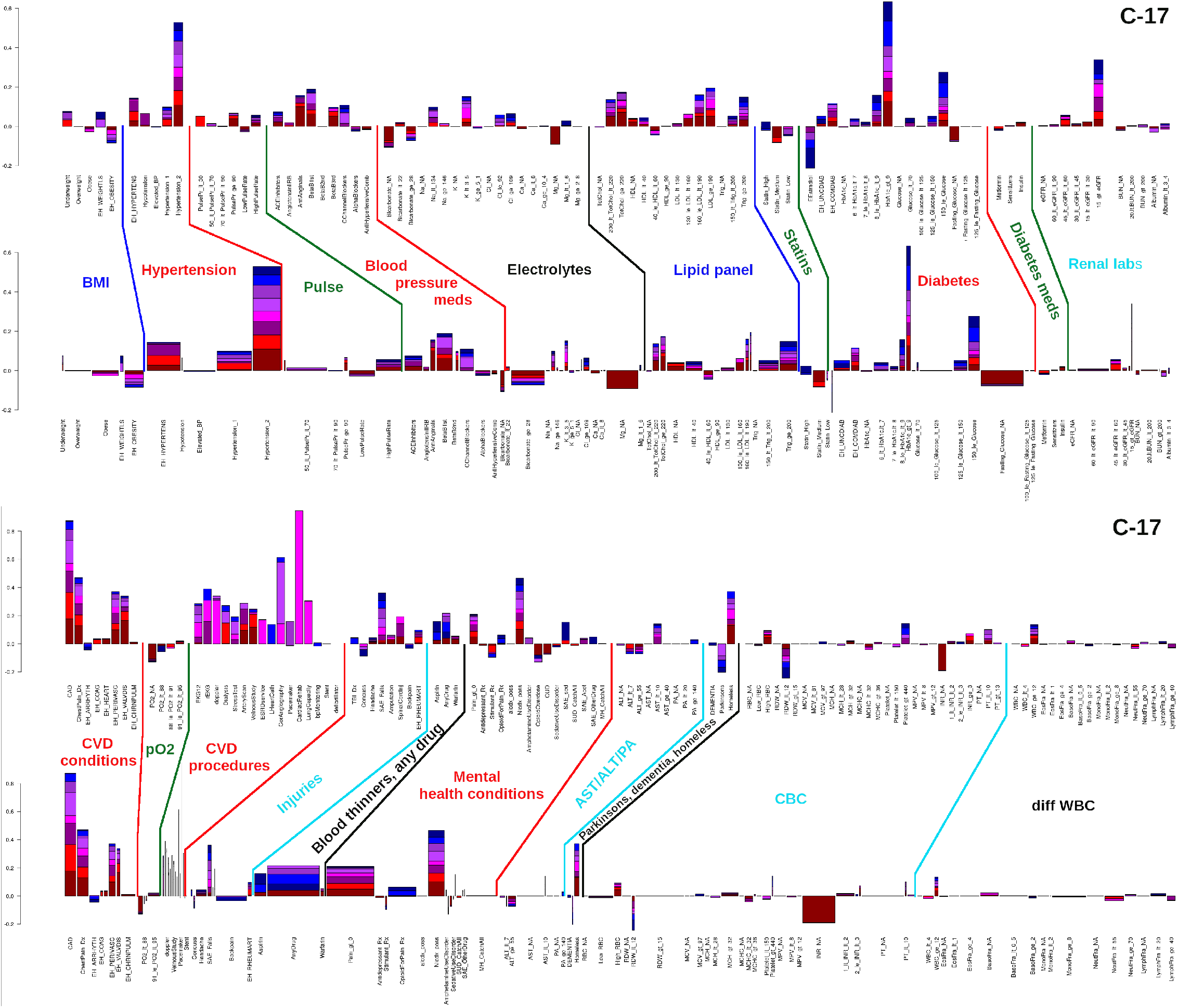
Stacked bar plots of model coefficients related to (top) metabolic syndrome, and (bottom) CVD disease progression, comorbidities, mental health, CBC, and dWBC, from the GLM model and C-17 cohort. Descriptions are the same as for Figs 4 and 5

**Figure S7:**
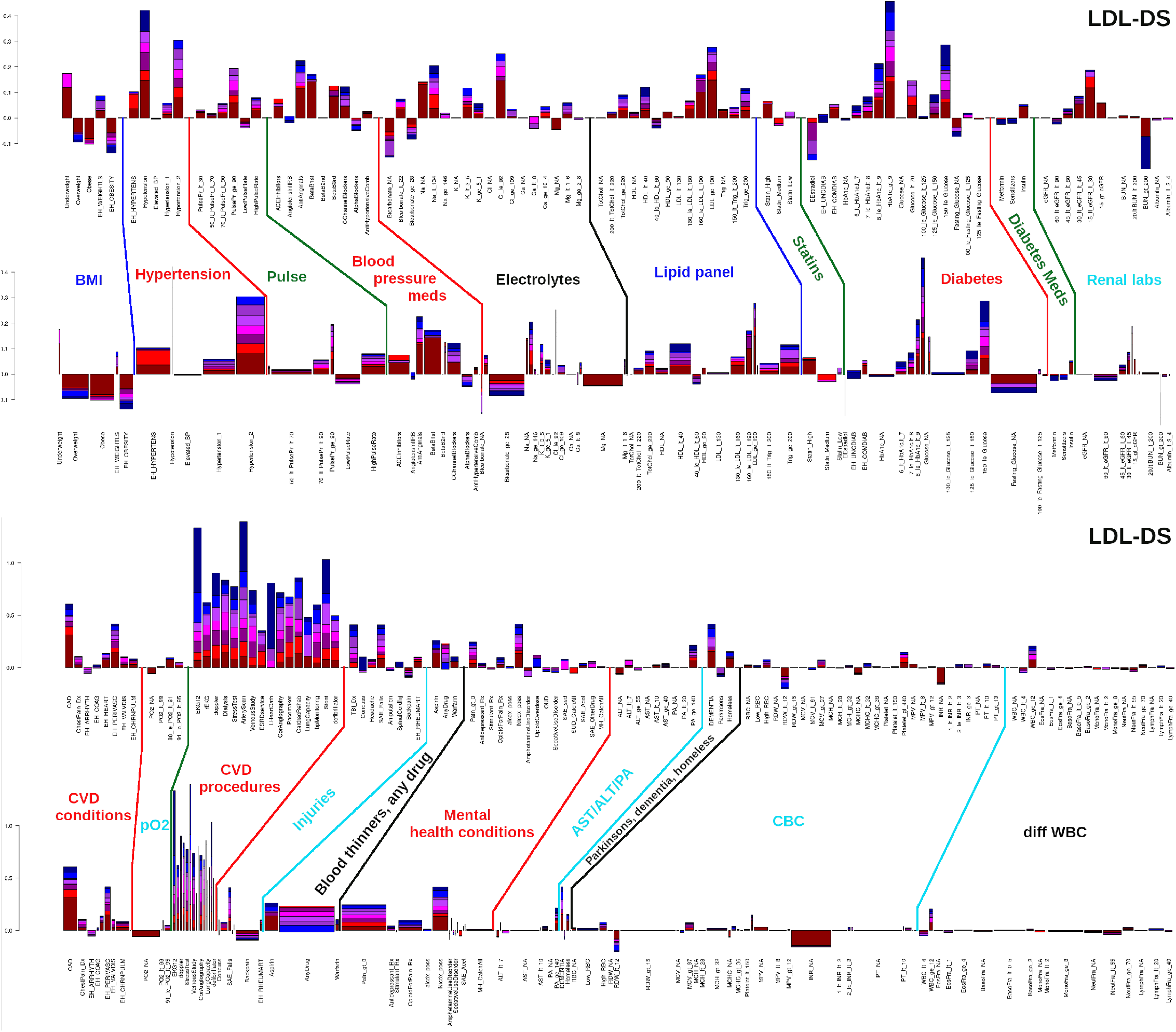
Stacked bar plots of model coefficients related to (top) metabolic syndrome, and (bottom) CVD disease progression, comorbidities, mental health, CBC, and dWBC, from the GLM model and LDL-DS cohort. Descriptions are the same as for Figs 4 and 5

**Figure S8:**
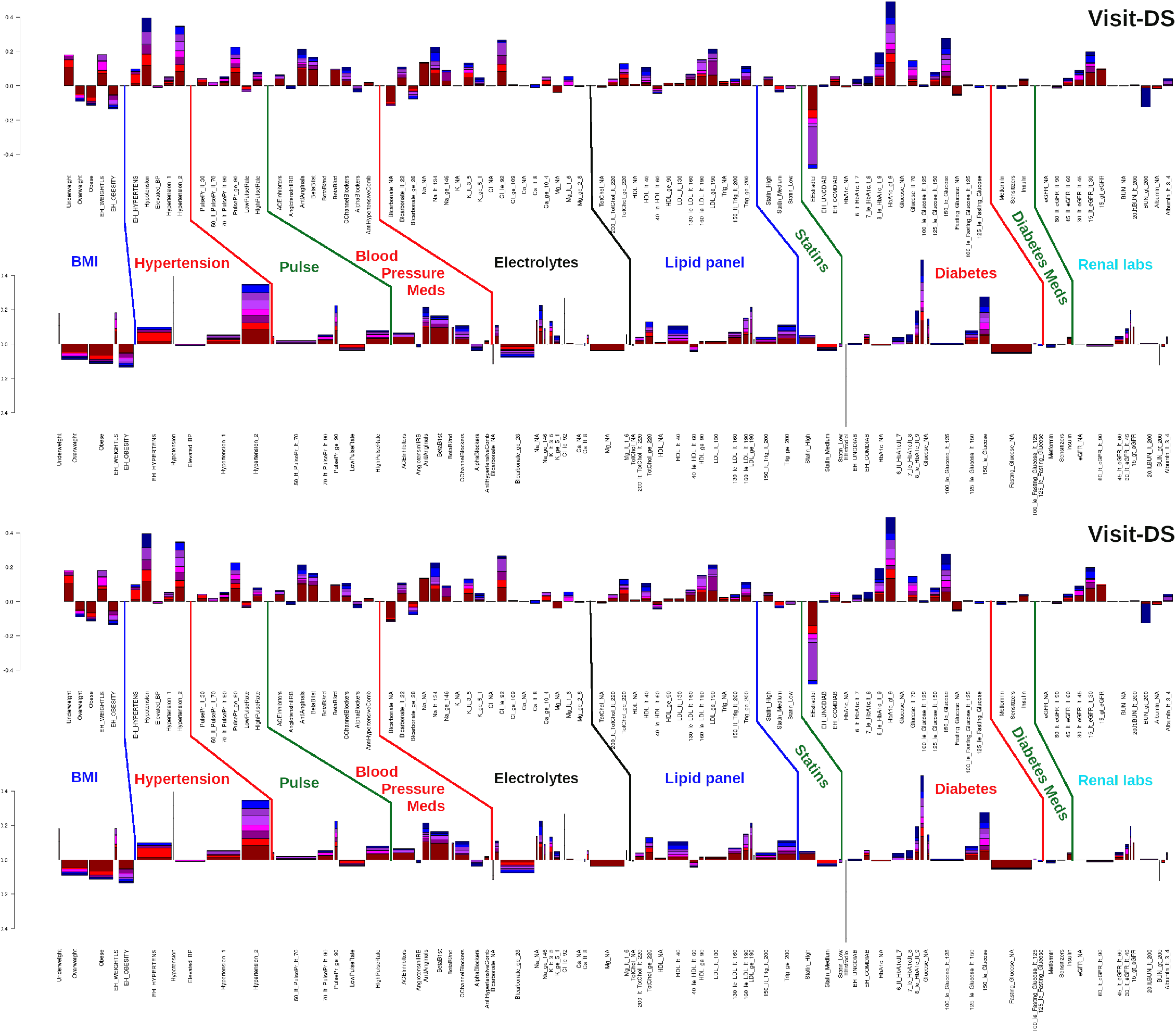
Stacked bar plots of model coefficients related to (top) metabolic syndrome, and (bottom) CVD disease progression, comorbidities, mental health, CBC, and dWBC, from the GLM model and Visit-DS cohort. Descriptions are the same as for Figs 4 and 5

Figure S9: Histograms of the number of patients with each Dx-encoded variable each month from 2000 through 2022. Also indicated are the top 10 most prevalent codes contributing to that variable’s signal, in the order of decreasing prevalence. ICD-9 and ICD-10 codes are interspersed for the purposes of ranking, and the ICD description is provided. Figures are provided as a separate pdf file with 47 pages, named CVD.pdf. These figures allow one to assess the utilization of the different codes through time and detect any discrepancies.

## Notes

### Competing Interest Statement

I have read the journal's policy and the authors of this manuscript have the following competing interests: CJO is an employee of Novartis Institute for Biomedical Research.

### Funding Statement

BHM and SD were awarded funding from the MVP Champion initiative, which was a Congressional allocation to VA & DOE, described at: https://www.energy.gov/articles/doe-and-va-team-improve-healthcare-veterans and administered through the Million Veteran Program at the VA ORD. The funders had no role in study design, data collection and analysis, decision to publish, or preparation of the manuscript.

### Author Declarations

This research was conducted with the written approval of the VA Central IRB with project number MVP014.

